# Quantifying the online news media coverage of the COVID-19 pandemic

**DOI:** 10.1101/2020.12.24.20248813

**Authors:** K. Krawczyk, T. Chelkowski, S. Mishra, D. Xifara, B. Gibert, D.J. Laydon, S. Flaxman, T. Mellan, V. Schwämmle, R. Röttger, J.T. Hadsund, S. Bhatt

**Affiliations:** Department of Mathematics and Computer Science, University of Southern Denmark, Odense, Denmark; Department of Management in the Network Society, Kozminski University, Warsaw, Poland; Department of Infectious Disease Epidemiology, Imperial College London, London, UK; Nupinion, London, UK; Department of Mathematics, Imperial College London, London, UK; Department of Biology and Biochemistry, University of Southern Denmark, Odense, Denmark

## Abstract

**Background:** Non-pharmaceutical interventions such as lockdowns, mask wearing and social distancing have been the primary measures to effectively combat the COVID-19 pandemic. Such measures are highly effective when there is strong population wide adherence which needs to be facilitated by information on the current risks posed by the pandemic alongside a clear exposition of the rules and guidelines in place. Here we address the issue of communication on the pandemic by offering data and analysis of online news media coverage of COVID-19.

**Methods:** We collected 26 million news articles from the front pages of 172 major online news sources in 11 countries (available at http://sciride.org). Using topic detection we identified COVID-19-related content to quantify the proportion of total coverage pandemic received in 2020. Sentiment analysis tool Vader was employed to stratify the emotional polarity of COVID-19 reporting. Further topic detection and sentiment analysis was performed on COVID-19 articles to reveal the leading themes in pandemic reporting and their respective emotional polarizations.

**Findings:** We find that COVID-19 coverage accounted for approximately 25% of all front-page online news articles between January and October 2020. Sentiment analysis of English-speaking sources reveals that the overall COVID-19 coverage cannot be simply classified as negative due to the disease subject matter, suggesting a wide heterogeneous reporting of the pandemic. Within this heterogenous coverage, 16% of COVID-19 news articles (or 4% of all English-speaking articles) can be classified as highly negatively polarized, citing issues such as *death, fear* or *crisis*.

**Interpretation:** The goal of pandemic public health communication is to increase understanding of distancing rules and maximize the impact of any governmental policy. Our results suggest an information overload in COVID-19 reporting that could risk obscuring effective policy communication. We hope that our data and analysis will inform health communication strategy to minimize the risks of COVID-19 while vaccination regimes are being introduced.

## 1. Introduction

The emergence of the novel coronavirus SARS-COV-2 and the resultant disease COVID-19 led to a pandemic that, to date, resulted in an estimated global mortality of 1.4 million people^1,2^. Due to the initial lack of the pharmaceutical measures targeting COVID-19, many governments resorted to unprecedented non-pharmaceutical interventions to control the spread of the pandemic^3,4^. It was estimated that introducing non-pharmaceutical preventative measures such as lockdowns, social distancing and mask-wearing had a significant effect on reducing the spread of COVID-19 and thus reducing the burden of the disease^5–8^. Therefore, in the absence of effective treatments or widespread rollout of vaccines, non-pharmaceutical interventions are still the primary mechanism available to control the spread of COVID-19^9^.

Crucial to the effectiveness of non-pharmaceutical interventions is their reliance on population wide adherence of governmental mandates, social distancing rules, stay-at-home orders and mask wearing by a substantial part of the society. This is highly dependent on the perception of the society towards such measures^9,10^. Such attitudes are invariably shaped and informed by the print and digital media, which allows one to build both an understanding of the changing disease landscape as well as the current prevention guidelines. As scientific information and evidence accumulates, it is expected that guidelines will shift and be clarified. In the digital age, one of the primary information sources for society are online news articles^11,12^. Effective communication via news articles on the current status of the pandemic and prevention guidelines can potentially affect how society responds to the non-pharmaceutical interventions introduced contributing to reducing the severity of the pandemic.

News articles have previously been shown as an effective way of tracking disease outbreaks, establishing their important role to study epidemics/pandemics in the digital age^13,14^. During the COVID-19 outbreak in particular, the role of news media in communicating the pandemic is crucial^15^. It was estimated that there were as many as 38 million English language articles on the coronavirus^16^. It was further demonstrated that there is a non-trivial amount of misinformation circulating on COVID-19 both on social media^17^ and in traditional news sources^16^. Navigating the information overload in an already complex patchwork of COVID-related topics adds to the burden in assuring societal literacy in understanding how to respond to the pandemic^18^.

An additional burden in responding to the pandemic is the emotional toll both the disease itself and the unavoidable distancing tactics have on the society ^19–21^. The sentiment of societal response to COVID-19 was gauged on social media^22–24^. All three studies of sentiments from COVID-19 conversations on social media showed a higher ratio of positive rather than negative emotions. In contrast, analysis of 25 English news media sources indicated that 52% of COVID-19 headlines evoked negative emotions^25^. News media have the power to shape societal adherence and behavior to the pandemic so extensive negative coverage or too much disparate information (information overload) could have detrimental effects to both the mental health of individuals and how effectively society responds to measures aimed at stemming the spread of the virus^26^.

At a time of global crisis, it is not surprising that there will be a lot of information pertaining to COVID-19 and that it would generally be negative. Quantifying the extent of both requires nuance as it is only then that one can draw meaningful conclusions on whether the coverage of the pandemic might be diverging from the goal of effectively communicating policy. Previous studies on the COVID-19 news articles showed that indeed there are many English-language articles on the topic^16^. Evenaga et al. sourced 38 million English-language COVID-19 articles by keyword search from LexisNexis that indexes 7 million sources. Quantifying the extent of COVID-19 information overload needs to take the amount of sources publishing data into account and thus must be calculated in context of all articles. Likewise, the sentiment of COVID-19 coverage also needs to be put in the context of overall negativity of consumed information. Aslam et al.^25^ analyzed 141,208 COVID-19 headlines showing that 52% of them carried negative sentiment. Similar results were reported by Chakraborty and Bose who collected COVID-19 news articles from GDELT (https://www.gdeltproject.org/) and found that pandemic coverage was mostly associated with negative sentiment polarization^27^. Neither of the studies contrasted COVID-19 sentiment distribution to the sentiment distribution of the sources they originated from. By contextualizing the sentiment distribution of pandemic reporting to this of the overall coverage it is possible to draw meaningful conclusions on whether the amount of COVID-19 information is indeed more negatively polarized than what news media consumers are exposed to.

Here, we address the issue of quantifying the extent of COVID-19 coverage and the sentiments that it might convey. We collected over 26 million articles from front pages of 172 major online news sources from 11 countries dating back to 2015 and compiled these into a re-usable database available at http://sciride.org. Firstly, we investigated trends in the subset of COVID-19 news with respect to all articles that appeared on the front pages in 2020. Secondly, we calculated the sentiment of COVID-19 articles and contrasted it to the background for each online news source and to other popular topics. Finally, we analyzed the leading sub-topics in COVID-19 coverage and assessed their sentiment polarization.

## 2 Methods

### 2.1 Curation of a database of front page news articles

In order to assess the extent of coverage and evoked sentiment of COVID-19 news we analyzed the landing pages from major Online News Sources (ONS) in countries with robust media presence. We selected the major ONSs from eleven countries: USA, UK, Canada, Australia, New Zealand, Ireland, Germany, France, Italy, Spain and Russia. We included an additional ‘International’ category to better reflect the global focus of certain ONSs.

For each country, the major ONSs were identified by reference to media profiles curated by the BBC and lists of news sites with most traffic, curated by SimilarWeb. Focusing on such ‘major’ national ONSs as defined by online visibility captures some of the main actors in shaping societal knowledge and opinions^28^. It should be noted that the focus on ONSs excludes the impact of social media, epistemic communities and other influences on public perception and is therefore limited. However, due to its depth of penetration, heterogeneity in political leanings, and reliability it represents a consensus that is strongly correlated with overall public perception.

For each ONS, we collected the archived front-page snapshots dating back to 2015 via a free service available through WebArchive (https://web.archive.org/), cutting off coverage in 2020 at 15th October. Each front page was then sourced for potential new items by a custom-based pipeline we developed for this project (see Supplementary Section 1). We defined each article as the combination of metadata elements *title* and *description*, that are akin to titles and abstracts in scientific publications^29^. Such metadata are reasonably standardized among ONSs and they offer a headline-like summarization of the article (typically designed for sharing on social media), making them suitable for topic detection and sentiment analysis. In total we collected 26,077,939 articles from front pages of 172 ONSs (Table 1) with the full list of contributing sources in Supplementary Table 1.

**Table 1.** Number of Online News source Sources and collected articles per country.

| **COUNTRY** | **NUMBER OF ONLINE NEWS SOURCES (ONS)** | **NUMBER OF ARTICLES** |
| --- | --- | --- |
| **CANADA** | 13 | 1,269,200 |
| **AUSTRALIA** | 8 | 1,124,859 |
| **ITALY** | 13 | 1,526,521 |
| **UK** | 21 | 4,977,792 |
| **USA** | 33 | 4,388,383 |
| **FRANCE** | 9 | 1,951,608 |
| **GERMANY** | 18 | 2,348,403 |
| **IRELAND** | 8 | 905,598 |
| **INTERNATIONAL** | 6 | 462,989 |
| **NEW ZEALAND** | 5 | 651,050 |
| **RUSSIA** | 19 | 3,348,825 |
| **SPAIN** | 19 | 3,122,711 |
| **TOTAL** | 172 | 26,077,939 |

We consider front pages as a reliable reflection of the information many users are exposed to whilst visiting these websites. This is opposed to other article collection strategies, such as RSS or downloading the entire website content, that have limited control over assessing how many people have actually read any given article^16,30^. Focusing our analysis efforts on articles from landing pages in major ONSs, gives us an opportunity to assess the number of COVID-19-related articles a large proportion of online news consumers might have been exposed to.

### 2.2 Topic Models

For each article we extracted from the front pages, we analyzed the text content of metadata title and description to determine whether it can be associated with one of the following topics: *CAT, SPORT, MERKEL, PUTIN, JOHNSON, BIDEN, TRUMP, CANCER, CLIMATE* or *COVID*. The non-COVID topics were selected as references with large expected volume of coverage (politicians) and those covering a wide range of sentiments (e.g. *CAT* as non-negative and *CANCER* as negative). Each of the topics was identified on the basis of keywords presented in Table 2. The only normalization we applied to the words for topic identification was case folding, otherwise the words were not stemmed nor lemmatized. Topics that were used solely for sentiment analysis, *CAT, SPORT, CLIMATE* and *CANCER* were not identified for non-English ONSs.

**Table 2.**
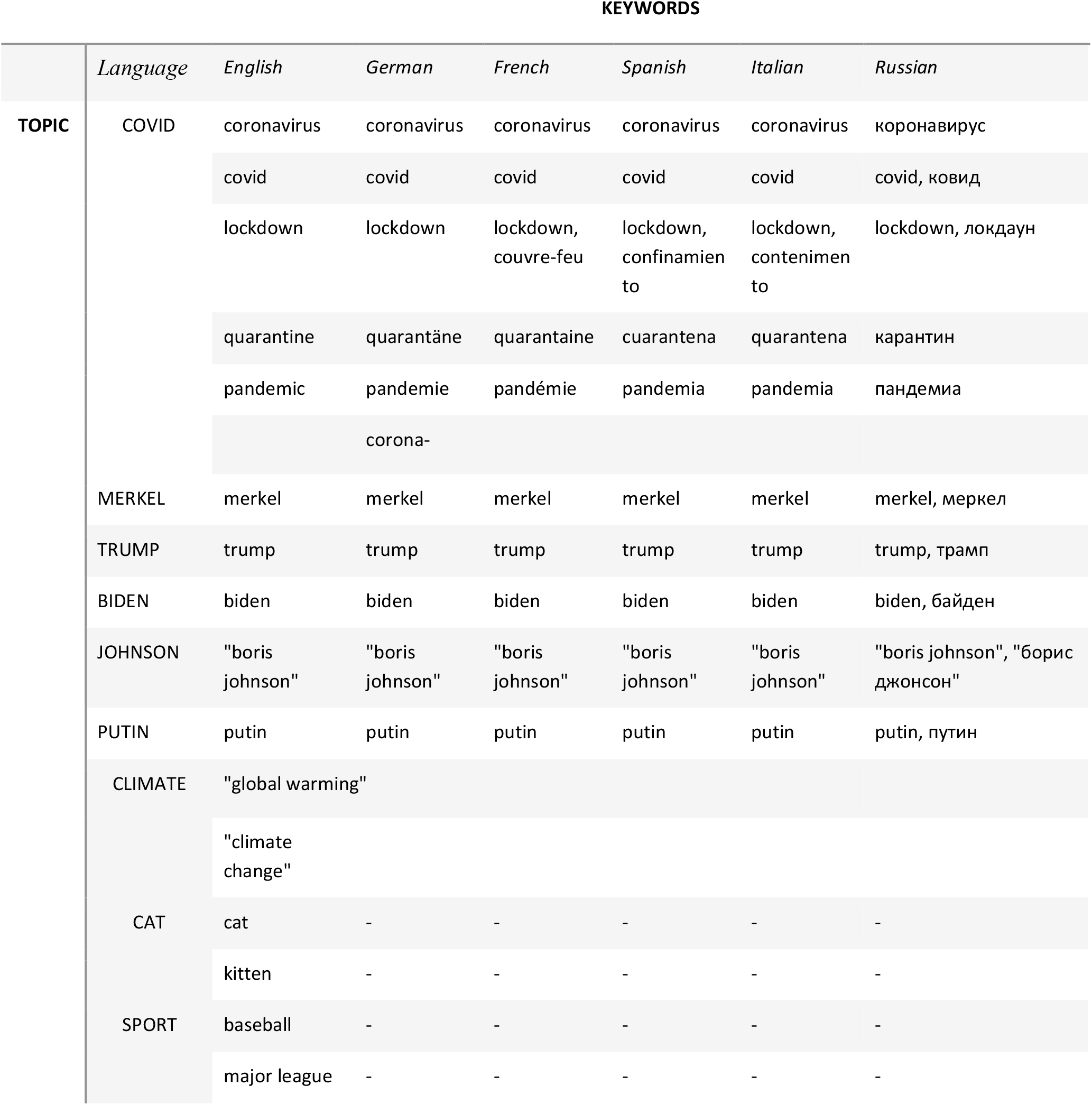
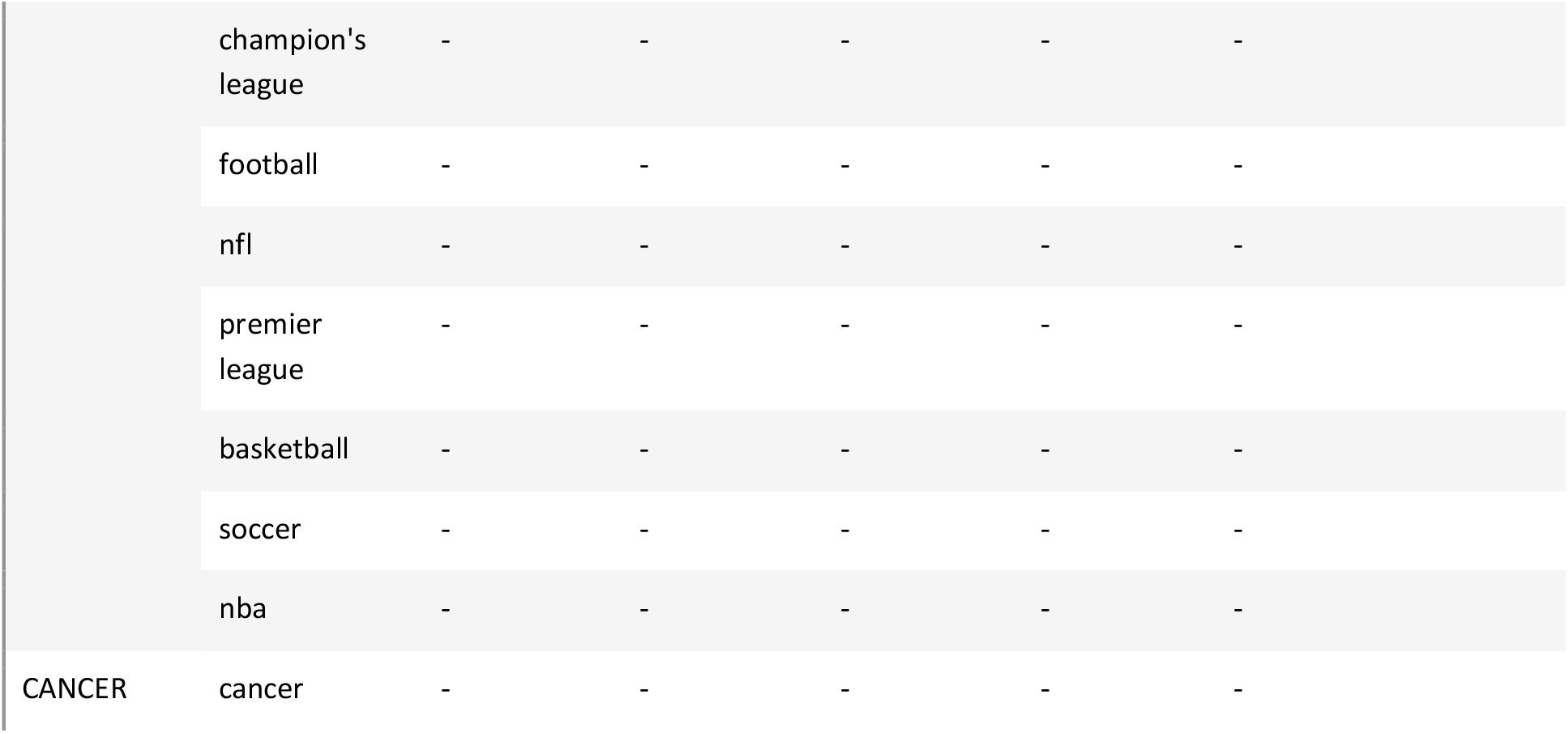
Keywords employed for topic detection. Topics *CAT, SPORT, CLIMATE* and *CANCER* were not identified in non-English speaking ONS as these were solely employed for sentiment analysis.

We further identified the sub-topics within COVID-19 coverage by a similar keyword-based approach. Since many of the subtopic words could have had several forms (e.g. dead, died, dies) we stemmed the words associated with each subtopic and present these in Table 3. An article was identified as pertaining to a subtopic if after stemming its title and description a token corresponding to a stemmed keyword in Table 3 was identified.

**Table 3.** COVID-19 news subtopics. Each of the subtopics was identified by the stemmed keywords (STEMMING).

| SUBTOPIC | STEMMING |
| --- | --- |
| CASE | case |
| CRISIS | crisi |
| DEATH | die, death |
| DISEASE | diseas |
| DISTANCING | distanc |
| FEAR | fear |
| HEALTH | health |
| HOME | home |
| HOSPITAL | hospit |
| INFECTION | infect |
| ISOLATION | isol |
| LOCKDOWN | lockdown |
| MASK | mask |
| OUTBREAK | outbreak |
| QUARANTINE | quarantin |
| SPREAD | spread |
| SYMPTOM | symptom |
| TEST | test |
| TREATMENT | treatment |
| VACCINE | vaccin |

### 2.3 Sentiment analysis of news articles using Vader

As the sentiment analysis measure, we employed Vader^31^. The Vader sentiment analysis tool is a well-established method to identify emotionally polarized content and it was previously applied to news articles. It is suitable for short snippets of text as in our titles and descriptions. For a given piece of text Vader provides a score between −1 and 1, with −1 being negative, 0 neutral and 1 positive. For instance “I find your lack of faith disturbing” offers Vader score of −0.42 whereas “I find your lack of faith encouraging” gives 0.5994. In our case a sentiment score for a single article consists of Vader compound score for the concatenation of article title and description.

Novel topics such as COVID-19 are associated with many subject-specific keywords and phrases (e.g. social distancing, lockdowns). Applying sentiment analysis to text containing such novel keywords is not guaranteed to produce desired results *a priori*. We assessed Vader sentiment annotations on articles identified as one of subtopics in Table 3. This revealed an artifact of the tool wherein ‘*positive coronavirus test*’ is labeled as emotionally polarised in the positive direction by virtue of the word *‘positive’*. In order to mitigate the effect of this subject-specific misclassification, the words *‘positive’* and *‘negative’* (for symmetry) were removed from articles related to coronavirus testing prior to applying Vader annotation.

### 2.4 Estimating topic polarization - Relative Sentiment Skew

Establishing whether coverage of a given topic is more negatively/positively emotionally polarized follows from comparing topic sentiment distribution to sentiment distribution of other articles not on that topic. Directly comparing topic sentiment distributions between different ONSs is not sound as sources can be more sensational/negative or toned-down/neutral giving radically different sentiment distributions. To address this issue we calculated whether specific topic coverage was more negative/positive/neutral relative to other articles in a specific ONS.

For each article ‘*a’* metadata title and description in English-language ONSs, we noted the Vader compound sentiment score, denoting it as *sent(a)*. For all 2020 articles from a given *TOPIC* and in a specific ONS, we calculated the mean Vader compound sentiment score (denoted *μ*_*ONS,TOPIC*_, eq. 1). As a reference statistic for distribution of sentiment scores not relating to *TOPIC*, we calculated the mean of articles from a given ONS that were not identified as a given topic (denoted 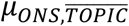, eq. 2).

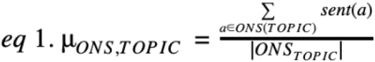

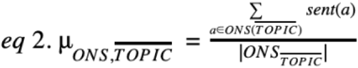

Where ONS is a particular Online News Source, *TOPIC* is one from Table 2 or Table 3, *ONS(TOPIC)* is the set of articles on a given topic from a particular ONS, and |*ONS*_*TOPIC*_| is the total number of articles on a given *TOPIC* in that ONS. A set of articles not identified as a given topic from a particular ONS is denoted as 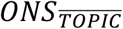.

For each topic in each ONS, we calculated the Relative Sentiment Skew (*rsskew*_*ONS,TOPIC*_) between topic mean sentiment and the mean of all other articles in the given ONS (eq. 3).

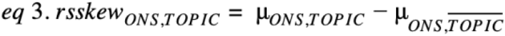

Relative Sentiment Skew is designed to indicate whether sentiment score distribution of coverage of any particular topic in a given ONS is more negatively (negative difference) or positively (positive difference) polarized as compared to other articles. Consider an example, where a topic has one positive (score +1) and two negative articles (score −1), and there are seven other non-topic articles that are all positive (score +1). In this instance our relative sentiment metric, *rsskew*_*ONS,TOPIC*_, is −1/3 - 7/7 = −1.33, which indicates greater negativity for the specific topic than other articles in the ONS. Note, we do not account for sample size variation as the denominator is generally very large (in thousands).

## 3. Results

### 3.1 Quarter of 2020 news was pandemic-related suggesting information overload

We estimated the extent of COVID-19 coverage in online news media by identifying articles relating to the pandemic and contrasting the number to the totality of articles between January and October 2020.

For each ONS, we performed topic detection categorizing each article title and description as relating to coronavirus (topic called *COVID*) if the title and description contained any keyword of a specific set. The keywords for this simplified topic detection model were chosen to maximize precision of content identification to avoid cross contamination with other topics. In English these keywords included *covid, coronavirus, lockdown, quarantine* and *pandemic*. The keywords were adjusted for the six languages that we used in this study: English, German, French, Spanish, Italian and Russian (Table 2).

For each ONS, we calculated the proportion of articles we detected to be on COVID-19 out of all the articles produced in a given ONS in 2020. Out of the total number of articles published in each of our 172 ONSs in 2020, we detect a mean of 25% (Figure 1) as COVID-19 related. Even though there exist certain variations between countries, the amount of volume is consistently large, with the lower bound at 20% and upper bound at 30%. Thus, even using our facile topic detection model we can demonstrate that 2020 online news coverage was dominated by COVID-19.

**Figure 1.**
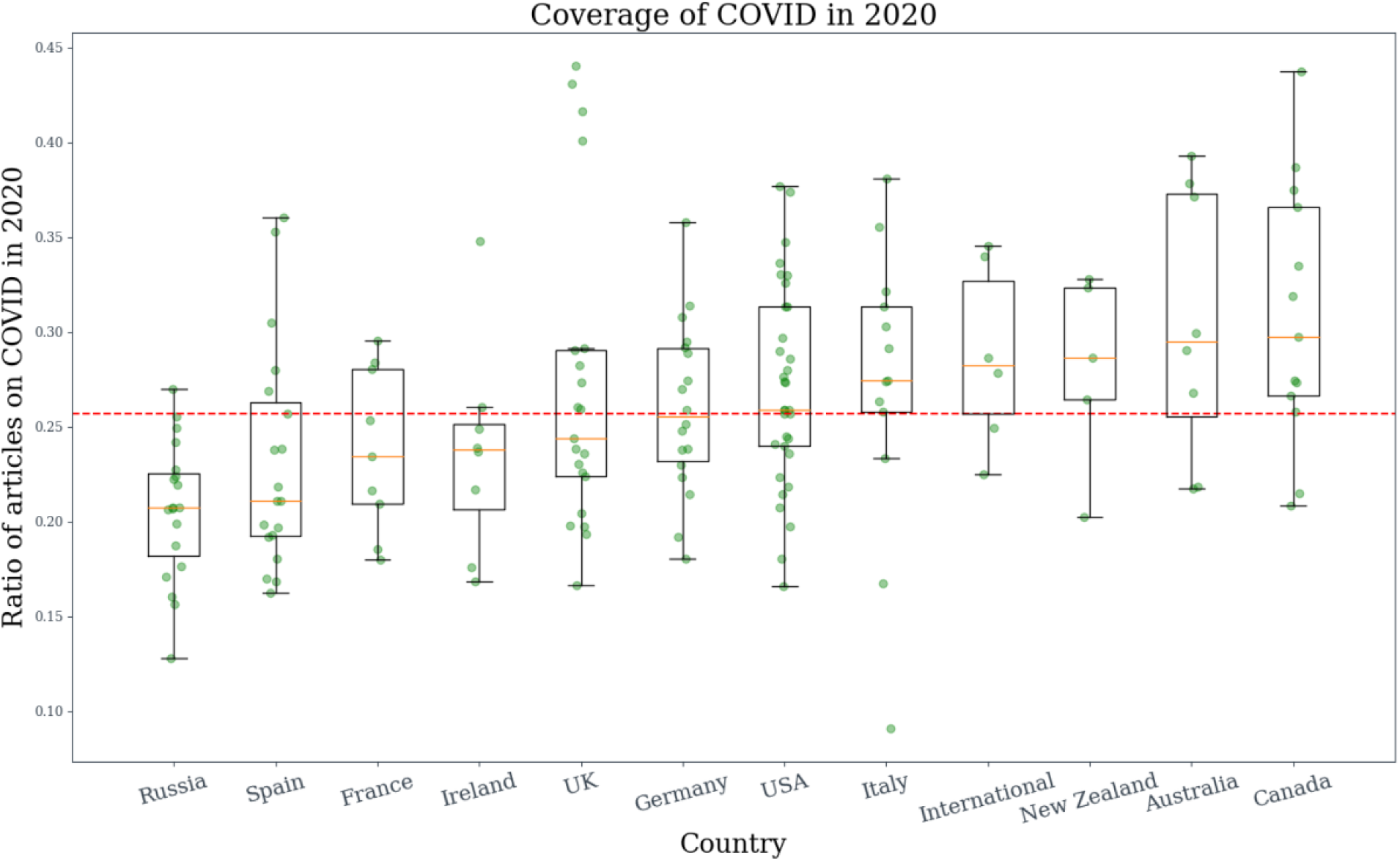
The extent of coronavirus coverage in 2020. We calculated the proportion of all COVID-19 articles as the proportion of all front page articles. The proportion was calculated for each Online News Source (ONS) separately and the proportions aggregated on national level. The green points represent the individual coverages of ONSs. The yellow line in each box represents the median with the upper and lower parts of the box the 75th and 25th percentiles respectively. The red dotted line indicates mean coverage across all ONSs.

In order to offer a point of reference to the overall coverage of COVID-19, we identified other global themes with regular and topical media presence (Table 2). We selected such topics as the politicians Donald Trump (*TRUMP*), Joe Biden (*BIDEN*), Boris Johnson (*JOHNSON*), Angela Merkel (*MERKEL*) and Vladimir Putin (*PUTIN*). Mentions of the different politicians are intuitively strongest in their home countries as demonstrated in Supplementary Figures 1-5 (e.g. Russia for Vladimir Putin). Nonetheless, each of the politicians received an order of magnitude less media attention than COVID-19 in 2020. Specifically, mean coverage of COVID-19 across 11 countries was 25% whereas politicians received 2.50% for *TRUMP*, 0.45% for *BIDEN*, 0.18% for *PUTIN*, 0.17% for *JOHNSON*, and 0.09% for *MERKEL*. Higher coverage of COVID-19 is also reflected on the national level. Even though 2020 was the election year in the USA, *TRUMP* as the sitting president received a mean 15.29% of 2020 media mentions in US ONSs as opposed to 25.91% mean for *COVID*. Furthermore, 2020 USA articles mentioning both *TRUMP* and *COVID* accounted for mean 3.82% coverage.

Our results provide a quantification of the extent of media attention received by COVID-19 that transcended different geographies and languages reflecting the global impact of the pandemic.

### 3.2 COVID-19 news sentiment analysis suggests heterogeneity of coverage

An important aspect in understanding how news coverage of COVID might affect societal responses to it, are the emotions that they would evoke^25^. To tackle this issue we performed sentiment analysis of the COVID-19 related news articles in 2020, contextualizing the sentiments to other reference topics and background sentiments for each ONS. This allows one to answer whether sentiment distribution of COVID-19 coverage was polarized as compared to what online news readers would be normally exposed to for a given ONS.

To quantify the emotional content of news article text, we employed a sentiment analysis tool, Vader, that had been previously applied to news article analysis^31,32^. For each ONS whose primary language was identified as English (91 out of 172), we created Vader annotations for both title and description of each of its articles. We grouped the articles by their annotated topics (Table 2) to offer a reference to the COVID sentiments. Topics representing the politicians (*MERKEL, TRUMP, JOHNSON, BIDEN* and *PUTIN*) were used as reference points for subjects with frequent coverage. We selected four additional topics so as to offer intuitive reference points on the positive/negative sentiment spectrum: *CAT, SPORT, CLIMATE* and *CANCER*. Topics of *CAT* and *SPORT* were used as a reference for topics that are not expected to be associated with negative sentiments. Likewise, topics of *CLIMATE* (identified by key-phrases *global warming, climate crisis*) and *CANCER* were used as references expected to be associated with negative emotions by virtue of their subject matter. Altogether, the individual sentiment annotations for each ONS were grouped by one of the topics: *CAT, SPORT, MERKEL, JOHNSON, BIDEN, TRUMP, COVID*, and *CANCER*.

For each ONS and each topic, we calculated the Relative Sentiment Skew (*rsskew*_*ONS,TOPIC*_, see methods) statistic that measures the polarization of a given *TOPIC* in a specific *ONS*. We noted how many topics had a negative or positive *rsskew* value and we present the numbers in Table 4. The skew in either positive or negative direction follows intuitive subject-matter allocations for the non-politician non-COVID topics (*CAT, SPORT, CANCER*), suggesting that they are appropriate references for assessing sentiment of COVID-19 articles on the positive-negative emotional spectrum. Though 74 out of 91 English-speaking ONSs (81.3%) hold general negative *rsskew* values in COVID-19 reporting, they are not substantially different from *rsskew=0* that indicates no polarization. The mean Relative Sentiment Skew for COVID is −0.04 with standard deviation 0.07 (Table 4). In particular, COVID-19 sentiment distribution is not as extreme as *CANCER* (100.0% per-ONS negative and mean *rsskew* −0.53), which should be the closest topic relative to COVID-19 by subject matter. In fact, the sentiment distribution for COVID-19 articles is more akin to coverage of *CLIMATE* (which was *a priori* expected to be negative akin to CANCER) or heterogenous subjects such as politicians (Figure 2).

**Table 4.** English-language ONS, having their *rsskew*_*ONS,TOPIC*_ of 2020 articles on a given topic either positive (>=0) or negative (<0). We had 91 English speaking ONSs in total, however in cases it was impossible to identify a certain topic in a given source, it was left out. The Relative Sentiment Skew column gives the mean *rsskew* values (μ) across the 91 ONSs together with the standard deviation (σ). The total number of articles we identified as a given topic across all ONSs is given in the column #Topic Articles.

| TOPIC | #POSITIVE ONS (%) | NEGATIVE ONS (%) | RELATIVE SENTIMENT SKEW | #TOPIC ARTICLES |
| --- | --- | --- | --- | --- |
| CAT | 64 (73.5%) | 23 (26.4%) | $\mu=0.12 \sigma=0.23$ | 2,746 |
| SPORT | 84 (92.3%) | 7 (7.6%) | $\mu=0.12 \sigma=0.08$ | 63,155 |
| BIDEN | 75 (83.3%) | 15 (16.6%) | $\mu=0.09 \sigma=0.11$ | 38,949 |
| JOHNSON | 57 (63.3%) | 33 (36.6%) | $\mu=0.04 \sigma=0.17$ | 22,613 |
| MERKEL | 38 (48.1%) | 41 (51.8%) | $\mu=-0.01 \sigma=0.25$ | 2,011 |
| COVID | 17 (18.6%) | 74 (81.3%) | $\mu=-0.04 \sigma=0.07$ | 589,701 |
| CLIMATE | 38 (41.7%) | 53 (58.2%) | $\mu=-0.04 \sigma=0.11$ | 7,195 |
| PUTIN | 33 (37.5%) | 55 (62.5%) | $\mu=-0.05 \sigma=0.23$ | 5,179 |
| TRUMP | 24 (26.3%) | 67 (73.6%) | $\mu=-0.06 \sigma=0.09$ | 157,702 |
| CANCER | 0 (0.0%) | 91 (100.0%) | $\mu=-0.53 \sigma=0.12$ | 9,548 |

**Figure 2.**
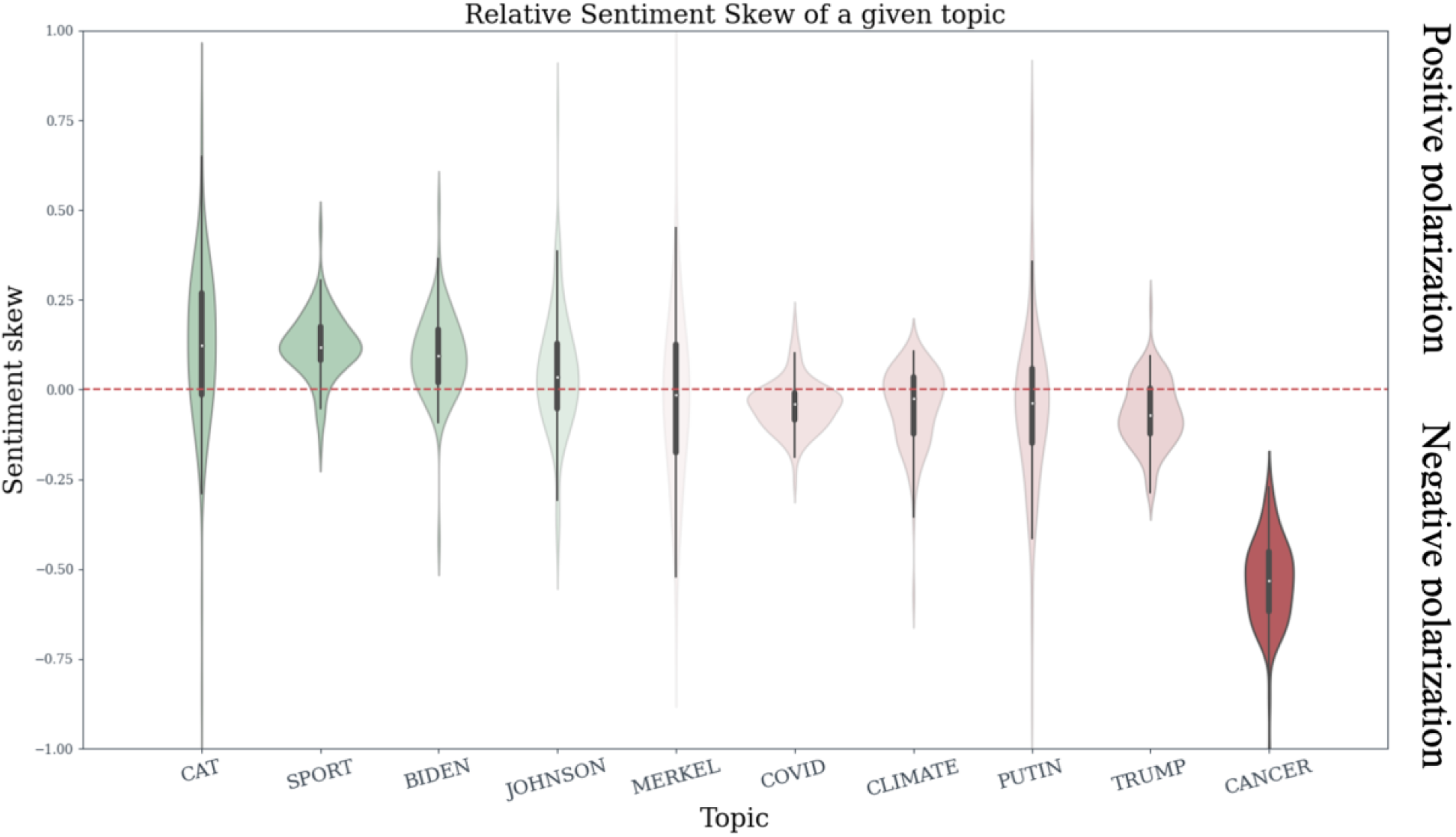
Relative Sentiment Skew of COVID-19 coverage. Each English-language ONS article title and description received Vader sentiment annotation between −1 and 1, being most negative and positive respectively. We noted the difference in the mean sentiment for a specific topic and mean sentiment for other 2020 articles in a given ONS (Relative Sentiment Skew, *rsskew*_*ONS,TOPIC*_, see methods). Relative Sentiment Skew in the negative direction indicated that news for the given topic were more negatively polarized than other articles and vice versa. The density of the Relative Sentiment Skew is plotted for each topic. Distributions are colored green if their *rsskew* was predominantly positive or red if it was negative (Table 4). Intensity of the color is scaled by the distance from the red dotted line at 0, indicating lack of difference between topic sentiment and all other articles in a given ONS.

These results suggest that the sentiment of COVID-19 coverage in online news media is quite heterogeneous, and certainly not as clearly polarized as *CANCER* (though volume of coverage might play a role, see Supplementary Section 3). Therefore COVID-19 articles cannot all be categorized as fully negative, contrary to expectation of pandemic subject matter. This suggests that coverage of COVID-19 was highly heterogeneous, with many themes contributing to the totality of messaging.

### 3.3 Highly sentiment-negative subtopics account for 16% of COVID-19 coverage, suggesting emotional pressure

We studied the text content of COVID-19 titles and descriptions metadata to reveal the leading themes associated with heterogeneous pandemic reporting.

To obtain an indication of the frequent subtopics, we calculated the most commonly used words and bigrams (combination of two consecutive words) to demonstrate the most frequent mentions in COVID-19 coverage. For each of the 91 English-speaking ONSs, we calculated the ranks of the single words and bigrams in articles that we identified as pertaining to COVID-19. The articles were further split within each ONS as *negative* (Vader score <-0.2, 247,542 articles), *positive* (Vader score >0.2, 192,643 articles) or *all* (any Vader score, 589,701 articles). Division into *positive, negative* and *all* aimed to reveal different keywords/bigrams that might have been mentioned based on the calculated sentiment. For one of the three subdivisions into *positive, negative* and *all*, we averaged the individual ONS ranks of each word and bigram found across all 91 ONSs. In Table 5 we present the top 20 words and bigrams that had the highest average ranks across all the 91 ONSs. Entries with stars in Table 5, indicate the words/bigrams that were among the top 20 only in a given sentiment subdivision (*positive, negative, all*). These words and bigrams reveal many themes that are intuitively associated with coronavirus pandemic such as *testing, vaccine, death* etc. In particular, negative articles however, appear to have unique top words and bigrams that are intuitively associated with negative emotions. In the singletons these are *death, crisi* and *fear* whether in bigrams these are *covid_crisi, covid_death, coronavirus_death* and *death_toll* (note that the words are stemmed).

**Table 5.**
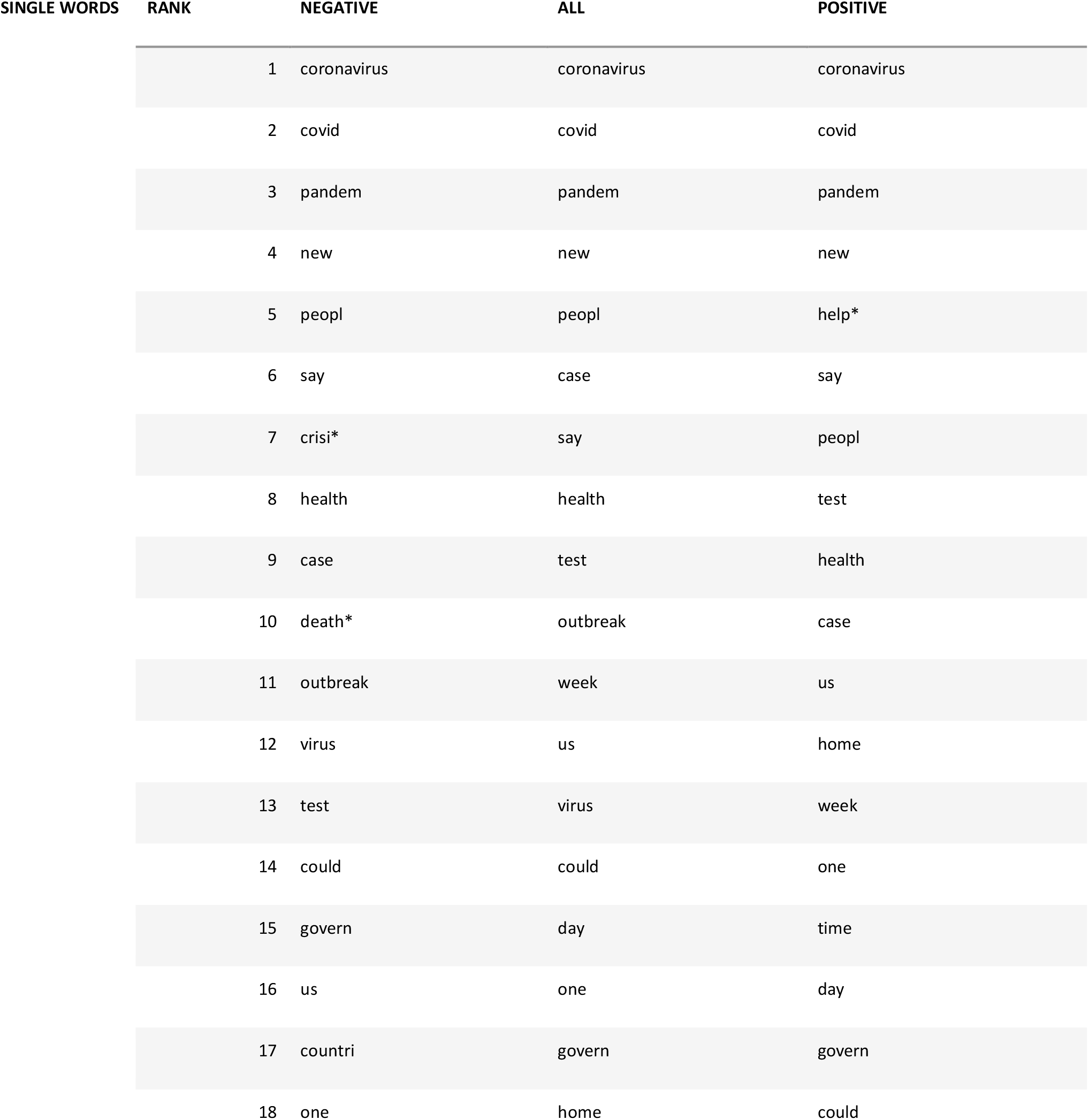
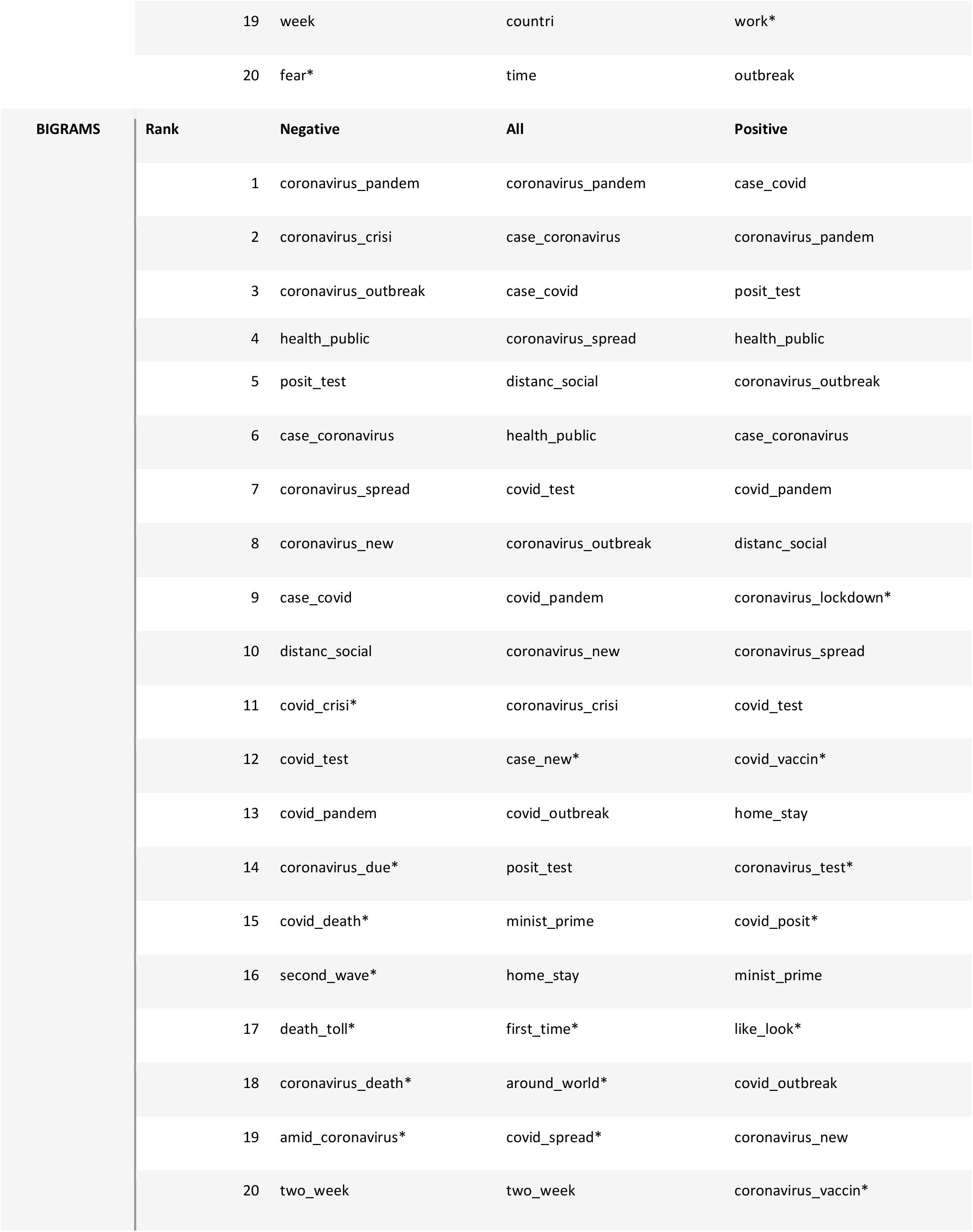
Top words and bigrams in English-speaking countries. For each of 91 English speaking ONSs, we calculated the most common word and bigrams and grouped these by Vader scores (>0.2 POSITIVE, <-0.2 NEGATIVE and any score for ALL). We averaged the ranks of words and bigrams across all ONSs and present the top-20 for each subdivision. Entries with starts (*) indicate elements that can be found in top-20 only in the specific subdivision into POSITIVE, ALL or NEGATIVE. All the words in the table are stemmed.

To calculate the news coverage proportion related to these top-themes, we created a constrained set of COVID-19 subtopics, based on the information from Table 5. We removed terms that pointed to wholesome coronavirus coverage such as *COVID, coronavirus, pandemic* or *news*. We extended the list of subtopics with ones that we did not find in Table 5 but considered as strongly related to COVID-19 coverage, such as *hospital, quarantine, symptom* or *isolation* with a full list of subtopics in Table 3. For each of the subtopics in Table 3, we calculated the per-ONS proportion of COVID coverage (Supplementary Figure 7) and per-ONS Relative Sentiment Skew for the subtopic (Supplementary Figure 8). The means of per-ONS coverage and sentiments are plotted against each other in Figure 3. The subtopics in Table 3 account for a mean of 67.14% of all COVID-19 articles across English speaking ONSs. Of these, top-three are *case, lockdown* and *death* that account for a mean of 9.29%, 8.56% and 8,08% of COVID-19 articles respectively. Figure 3 and Supplementary Figure 8 suggests that out of *case, death* and *lockdown*, only *death* carries firmly polarized sentiment, with *case* and *lockdown* not being significantly skewed in either positive or negative direction. Figure 3 reveals that of all the subtopics, *fear, crisis* and *death* indicate a substantial polarization towards an extreme emotion, intuitively for all three, towards negative.

**Figure 3.**
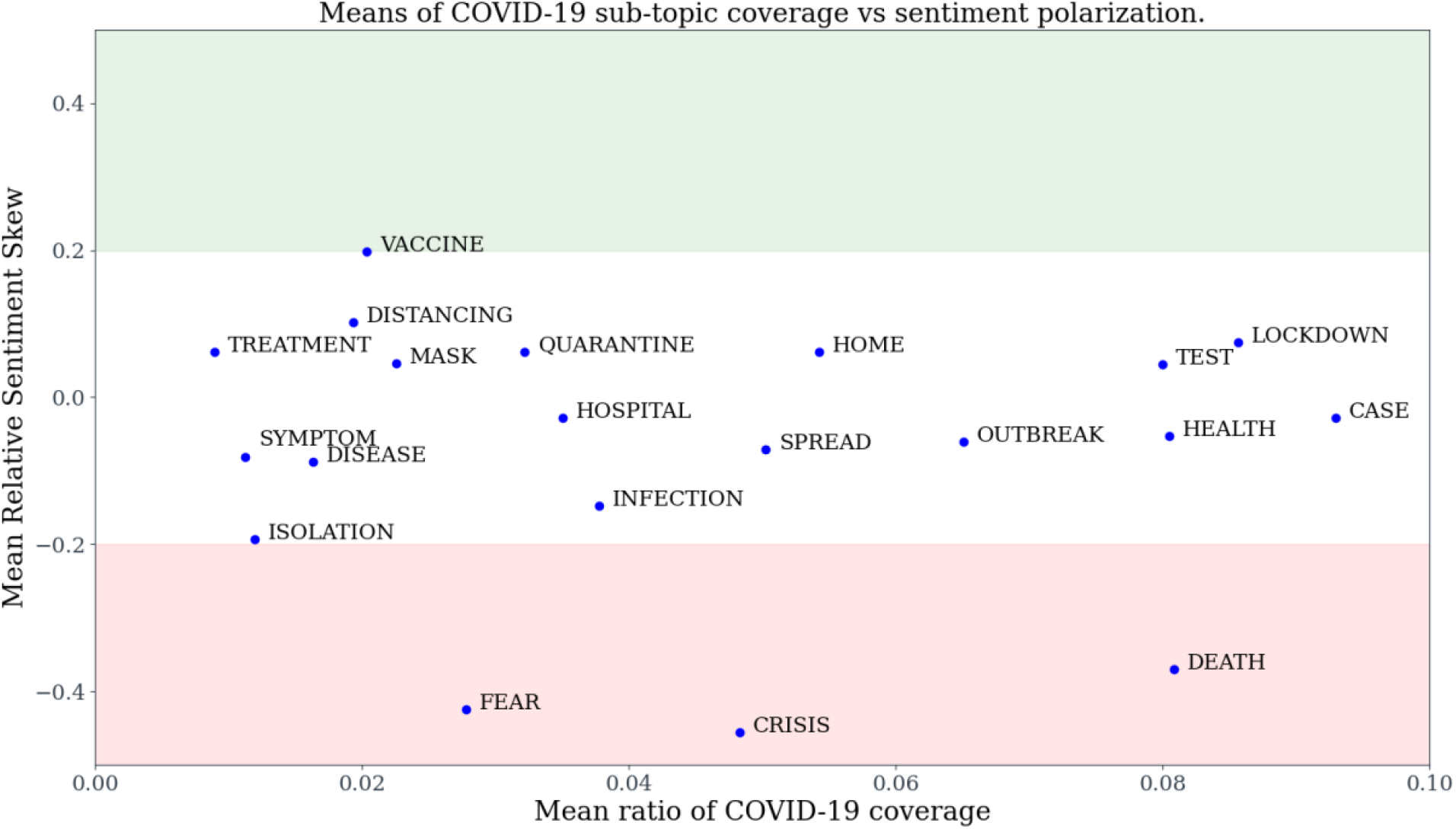
COVID-19 subtopic coverage and sentiment means. We calculated the mean coverage of each subtopic and mean Relative Sentiment Skew of each subtopic. Coverage is expressed as the mean of ratios of subtopic in a given ONS against all COVID-19 articles in the same ONS. Sentiment is the mean of subtopic Relative Sentiment Skew for all ONSs. The shaded areas illustrate regions with Relative Sentiment Skew above 0.2 (green), between 0.2 and −0.2 (white), and below −0,2 (green).

We analyzed the impact on news sentiment in 2020 of the three most negative subtopics, *fear, crisis* and *death* (Figure 3). For each ONS we calculated the mean sentiment of all the 2020 articles after removing those COVID-19 articles that mentioned *fear, crisis* or *death*. For all 91 ONSs removing the COVID-19 articles mentioning *fear, crisis* or *death* resulted in a statistically significant shift towards mean positive sentiment (see Supplementary Section 5). By contrast, mean sentiment of 2020 articles after removing all the sentiment-heterogeneous (Figure 2) COVID-19 articles results in significant shift towards positive in 40 out of 91 (43.95%) ONSs, for 11 out of 91 (12.08%) ONSs a significant shift towards the negative and no statistically significant result for 39 out of 91 (42.85%) ONSs. Altogether articles mentioning *fear, crisis* and *death* account for mean 16% of our COVID-19 articles and due to their highly-polarizing nature and relative volume might play a significant role in shaping the societal perception of the pandemic.

Of the three most negative topics of *fear, crisis* or *death*, the latter is the most frequently mentioned in the context of COVID-19 (Figure 3). In total, death mentions in the context of COVID-19 accounted for a mean of 2.33% of all coverage in 91 English-speaking ONSs. All death mentions in 2020 in 91 English-speaking ONSs account for mean 5.74% coverage whereas in the pre-COVID period of 2015-2019 they accounted for 4.07%. Therefore, we can identify that death in the context of COVID-19 constituted a significant proportion of negatively-associated coverage that appears to have contributed to overall death reporting in the news.

These results demonstrate that even though the overall coverage of COVID-19 in 2020 is not significantly sentiment-polarized, there is a non-trivial proportion of negative news that contribute to overall reporting negativity in 2020.

## 4. Discussion

Non-pharmaceutical interventions are drastic measures that reduce the pandemic casualties before widespread vaccinations and/or treatment regimes become norm. Such methods however are not effective without societal adherence. Information received by a population in times of a pandemic shapes collective adherence to policies introduced to stem its spread. Currently the internet is the main source of health information for many people in developed countries^33^.

Comprehensive analysis of COVID-19 information received by a population would require thorough analysis of the possible internet news sources and the user exposure to each piece of information received. Online ecosystem is extremely heterogeneous with information discovery channels spread across traditional news sites, blogs, social media and many others. Within each of such platforms, information itself can take different forms (e.g. text length, format). How users interact with the information also has a great effect on the amount of attention a given piece of information receives (e.g. extent of sharing on social media or more visible position on a website). Analysis of COVID-19 information from all the different online sources is not tractable. Therefore in our study of COVID-19 news, we focused on articles published directly on landing pages of major news sites as they offered a good balance between the proportion of online traffic they captured and format standardization being favorable for text analysis.

Direct access to major news sites accounts for 76% of media consumption online^28^. Taking into account the landing pages of such sources implicitly captured articles that might have been seen by online users. Thus, our analysis of content from front pages of major news sites should encompass a significant proportion of sources shaping knowledge of the pandemic covering reporting across different languages and geographies. In total, we curated a dataset of 26 million articles from 172 major web-traffic generating ONSs in 11 countries.

Out of the articles we collected, we identified COVID-relevant documents as well as a selection of other topics to act as intuitive references for coverage volume and sentiment analysis. Rather than analyzing the full-body of each article we analyzed the metadata titles and descriptions where the main subject matter can be expected to be referenced. We employed a facile topic identification method, using a limited set of keyword mentions. The choice of keywords used was selected so as to make it unlikely that an article does not make the corresponding topic its subject matter if it were referenced in metadata title and description. This avoided the caveat of tangential references to certain topics mentioned in the full article body or ambiguities that might arise by using more sophisticated topic modeling algorithms^34^. Unlike more complex topic modeling methods, our approach did not capture much more subtle references to these topics, underestimating the total coverage.

Nonetheless, even using our very simple approach to topic modeling of COVID-19, we still identified a non-trivial amount of articles on the front pages of our ONSs referring to it. We estimate that a mean of 25% of our sample of front-page articles from 11 countries in 2020 mention COVID-19 in their titles and descriptions. Our method had reduced topic identification recall by not accounting for more subtle references to COVID-19 and the totality of the articles was certainly contaminated by retrieval of erroneous links that were not actual news articles. Therefore, the factual proportion of articles on news front pages referencing COVID might have been indeed higher. We envision that the amount of reporting on a topic of general interest like COVID-19 needs to be balanced. Too little information might leave the population under-informed and ill-equipped to respond appropriately. Too much coverage on the other hand runs the risk of obscuring information that is crucial for individuals to build an understanding of the pandemic and how to act in order to stay safe.

Reporting on the pandemic cannot be perceived only for its informative function. It is unknown what effect a constant reporting on the cases, casualties and containment methods could have on adherence to distancing rules or mental health^19,35–38^. We employed sentiment analysis to study the large volume of identified COVID-19 articles by analyzing emotions they could evoke. Though current sentiment analysis methods fall short of identifying complex nuance, they offer a good approximation for the position of text on the emotional spectrum (negative, neutral, positive). Employing sentiment analysis we found that overall COVID-19 reporting was not markedly polarized in either positive or negative direction, as opposed to another health reference topic such as cancer. It is contrary to what might be expected by virtue of the pandemic subject matter, suggesting heterogeneity in reporting. Such heterogeneity might have come about by the ever-permeating nature of the pandemic, acting as a background to much activity in 2020. Nevertheless, we found that negatively-polarized COVID-19 articles mentioning *death, fear* and *crisis*, account for 16% of pandemic articles, with death being most widely referenced. Such non-trivial volume of negatively-associated articles significantly skews the sentiment of 2020 reporting in the negative direction.

Altogether our results offer a quantification of COVID-19 reporting that substantiates widespread qualitative observations (e.g. pandemic received an unprecedented amount of media attention). This data can offer insight for shaping the discussion on health communication with the public to maximize the effect of policy introduced to stem the spread of the pandemic. Our retrospective analysis of health communication on the pandemic during the first two waves indicates signs of information and emotional overload that might have obscured understanding of policy. We hope that our findings could inform how to adjust the communication so as to minimize the risk of the third wave whilst vaccination regimes are being introduced.

## Data Availability

We make the collected online news article data available via our website at http://sciride.org.

## Contributors

KK and SB conceived of and designed the study. KK and TC extracted the data. KK, TC and SM had access to and verified the data. KK, TC and SM processed the data. All authors contributed to the analysis and writing of the manuscript. All authors revised the manuscript.

## Acknowledgements

SB would like to acknowledge the BMGF, the Academy of Medical Sciences, The NIHR BRC and the UKRI. We would also like to acknowledge Microsoft AI for health and Amazon AWS for computing resources.

## Data Sharing

We make the collected online news article data available via our website at http://sciride.org.

