## Supplementary for "Quantifying the online news media coverage of the COVID-19 pandemic"

### Supplementary Material for *Quantifying the online news media coverage of the COVID-19 pandemic.*

### Supplementary Section 1. Data retrieval.

We retrieved news articles from major international and national online news sources (ONS) in USA, UK, Canada, New Zealand, Australia, Ireland, Russia, Italy, France, Spain and Germany. We selected the ONS in each country by examining the top news providers calculated by website traffic according to SimilarWeb (http://similarweb.com) metrics and those that were listed under the BBC media profiles. We were able to confirm internally that numbers from SimilarWeb were accurate by comparing their web traffic estimates to Google Analytics traffic from a major website. This approach to select our media sources increases the likelihood that our ONS would capture a significant portion of the news consumption within each country.

For each ONS we retrieved the landing page snapshots from WebArchive captured since 2015. WebArchive is a reliable online service that specializes in archiving online web content for 477 billion webpages. Since WebArchive frequently captures website snapshots (up to several times per day), we considered it a reliable indicator of the evolution of the news content on each of our ONSs.

For each ONS front-page snapshot extracted from WebArchive we extracted all the links which stayed within the domain of the ONS (see Supplementary Table 1). For each of the links we extracted the raw html, if it was possible directly from the source, otherwise the archived version from WebArchive. If it was impossible to retrieve the link contents either from the original source or the WebArchive, we flagged such links as 'retrieval error'. The errors in retrieval were defined as pages where the server did not respond correctly, the webpage was unavailable (e.g. 404 errors) or if access to certain resources was blocked (e.g. subscription needed). We curated a global list of error messages after examining the responses for each ONS. In order to assure that we obtain a representative reflection of each ONS, we set the retrieval error rate at 6% (which allowed a good balance between including many ONSs and reliability of link retrieval). There were certain exceptions to this rule where we noted that the links we could not retrieve could not possibly have been article related (e.g. repubblica.it, stuff.co.nz). The error rates in retrieval for each ONS are given in Supplementary Table 1.

For each raw html page we extracted the metadata called 'title' and 'description'. Such data elements are typically available for news article content as, among others, they allow for condensed snippets of text to be shared on social networks such as Twitter and Facebook. They are akin in spirit to titles and abstracts of scientific information and serve a similar function of conveying the essence of the document. All things considered, title and description metadata offer the advantage of being standardized for retrieval, their text length and the condensed information between otherwise heterogenous for each ONS.

For each ONS, the titles and descriptions were concatenated and associated with all the links pointing to the same text content. This allowed us to collapse url-distinct links pointing to the same content. Such normalization reduces the number of multiple-counts for each article as the raw URL might be updated over several days, or there could be links to comments or video sections all referencing the same core content. The number of such unique links are given in the ‘Articles’ column in Supplementary Table 1. Furthermore, we analyzed the titles and descriptions associated with the greatest number of links. This allowed us to identify ONS-specific error pages since multiple links that cannot be retrieved linked to the same error title page. Such information was fed backwards into our list of error responses, iteratively refining the retrieval procedure.

Retrieval of all the links from ONS meant that we captured many non-article structural links, such as category pages, terms of use etc. Each title and description text pair was given a per day count as well as count of how many times links pointing to it were observed in all the WebArchive snapshots. Pre-2020 articles provided a large dataset of counts of links that appear very often on ONS front pages, indicating sections etc. Title and description text pairs with per day count above 100 were discarded from the analysis as these were deemed to be structural non-article links.

As the final article filter for each ONS, we performed language classification using the Python library langdetect. Each ONS was associated with a language that articles are expected to be written in (e.g. English for FoxNews and German for Der Spiegel). Articles whose titles and descriptions were not classified as the main language of the ONS were discarded from the analysis. Structural website elements (e.g. section links) or erroneous text was typically language-misclassified (e.g. Daily Telegraph) further reducing the contamination of our dataset with non-article content. In certain cases, such as the BBC, a great number of links were in fact not in English, which is due to the global reach of the service. Certain USA ONS had a sizable proportion of articles in Spanish (e.g. Los Angeles Times or Chicago Tribune). We considered such articles tangential to the overall coverage of these ONS and decided not to include these in the analysis.

The raw front pages, raw individual articles and processed articles are available for download via http://sciride.org.

**Supplementary Table 1.** Statistics of retrieval for news media domains. For each domain, we report the number of unique links we were able to identify in the front pages (total links). Out of those links we report how many produced retrieval errors (Number of bad links) and the percentage thereof (%bad links). The Articles column reports the total number of unique articles we extracted from each domain. The Rejected column reports the number of articles that were rejected as being unlikely news articles.

| DOMAIN | COUNTRY | TOTAL LINKS | NUMBER OF BAD LINKS | %BAD LINKS | ARTICLES | REJECTED |
|---|---|---|---|---|---|---|
| **CTVNEWS.CA** | Canada | 231,051 | 99 | 0 | 144,582 | 103 |
| **THEGLOBEANDMAIL.COM** | Canada | 196,001 | 995 | 0.5 | 181,441 | 222 |
| **LAPRESSE.CA** | Canada | 169,789 | 2,259 | 1.3 | 153,650 | 237 |
| **GLOBALNEWS.CA** | Canada | 150,027 | 105 | 0 | 130,438 | 324 |
| **ICI.RADIO-CANADA.CA** | Canada | 147,066 | 1,613 | 1 | 104,264 | 524 |
| **THESTAR.COM** | Canada | 140,995 | 417 | 0.2 | 134,238 | 160 |
| **MONTREALGAZETTE.COM** | Canada | 123,027 | 4,818 | 3.9 | 112,947 | 330 |
| **CBC.CA** | Canada | 103,159 | 662 | 0.6 | 94,849 | 128 |
| **TORONTOSUN.COM** | Canada | 101,807 | 1,129 | 1.1 | 87,497 | 340 |
| **VANCOUVERSUN.COM** | Canada | 72,337 | 2,292 | 3.1 | 65,791 | 243 |
| **JOURNALDEMONTREAL.COM** | Canada | 39,024 | 155 | 0.3 | 35,802 | 123 |
| **MACLEANS.CA** | Canada | 17,890 | 72 | 0.4 | 17,040 | 158 |
| **RCINET.CA** | Canada | 11,558 | 59 | 0.5 | 6,661 | 4,032 |

| | | | | | | |
|---|---|---|---|---|---|---|
| **TOTAL** | Canada | 1,503,731 | - | - | 1,269,200 | - |
| **NEWS.COM.AU** | Australia | 273,231 | 2,405 | 0.8 | 247,873 | 203 |
| **SMH.COM.AU** | Australia | 250,848 | 3,926 | 1.5 | 227,109 | 562 |
| **THEAGE.COM.AU** | Australia | 233,318 | 4,043 | 1.7 | 211,739 | 393 |
| **ABC.NET.AU** | Australia | 177,060 | 5,205 | 2.9 | 149,086 | 153 |
| **AFR.COM** | Australia | 124,697 | 517 | 0.4 | 116,897 | 482 |
| **9NEWS.COM.AU** | Australia | 82,263 | 1,163 | 1.4 | 66,585 | 145 |
| **THEWEST.COM.AU** | Australia | 57,363 | 208 | 0.3 | 56,774 | 132 |
| **SBS.COM.AU** | Australia | 56,958 | 822 | 1.4 | 48,796 | 196 |
| **TOTAL** | Australia | 1,255,738 | - | - | 1,124,859 | - |
| **REPUBBLICA.IT** | Italy | 376,716 | 44,125 | 11.7 | 193,065 | 771 |
| **ILSOLE24ORE.COM** | Italy | 211,950 | 5,777 | 2.7 | 133,605 | 1,885 |
| **LASTAMPA.IT** | Italy | 191,831 | 1,272 | 0.6 | 172,802 | 1,355 |
| **HUFFINGTONPOST.IT** | Italy | 188,637 | 616 | 0.3 | 106,988 | 625 |
| **ILFATTOQUOTIDIANO.IT** | Italy | 183,423 | 551 | 0.3 | 100,594 | 304 |
| **ANSA.IT** | Italy | 178,569 | 288 | 0.1 | 169,341 | 2,994 |
| **ILMESSAGGERO.IT** | Italy | 174,755 | 1,065 | 0.6 | 158,998 | 174 |
| **ILMATTINO.IT** | Italy | 163,282 | 386 | 0.2 | 129,564 | 292 |

| | | | | | | |
|---|---|---|---|---|---|---|
| **CORRIERE.IT** | Italy | 133,387 | 974 | 0.7 | 105,816 | 687 |
| **TISCALI.IT** | Italy | 102,778 | 2,512 | 2.4 | 93,442 | 404 |
| **LIBEROQUOTIDIANO.IT** | Italy | 86,152 | 5,111 | 5.9 | 66,135 | 126 |
| **FANPAGE.IT** | Italy | 81,871 | 199 | 0.2 | 80,235 | 302 |
| **RAI.IT** | Italy | 17,892 | 511 | 2.8 | 15,936 | 377 |
| **TOTAL** | Italy | 2,091,243 | - | - | 1,526,521 | - |
| **DAILYMAIL.CO.UK** | UK | 1,529,517 | 49 | 0 | 829,047 | 86 |
| **MIRROR.CO.UK** | UK | 859,725 | 1,087 | 0.1 | 468,632 | 603 |
| **THESUN.CO.UK** | UK | 791,669 | 985 | 0.1 | 509,583 | 185 |
| **REUTERS.COM** | UK | 543,163 | 7,725 | 1.4 | 328,651 | 3,288 |
| **THETIMES.CO.UK** | UK | 486,590 | 256 | 0 | 415,801 | 65 |
| **DAILYSTAR.CO.UK** | UK | 416,041 | 226 | 0 | 299,874 | 287 |
| **EXPRESS.CO.UK** | UK | 403,722 | 385 | 0 | 401,076 | 172 |
| **TELEGRAPH.CO.UK** | UK | 319,967 | 625 | 0.1 | 278,797 | 2,585 |
| **THEGUARDIAN.COM** | UK | 290,741 | 45 | 0 | 288,567 | 304 |
| **LIVERPOOLECHO.CO.UK** | UK | 222,963 | 288 | 0.1 | 118,811 | 261 |
| **BBC.COM** | UK | 215,915 | 137 | 0 | 163,727 | 26,561 |
| **INDEPENDENT.CO.UK** | UK | 208,125 | 226 | 0.1 | 205,206 | 281 |

| | | | | | | |
|---|---|---|---|---|---|---|
| **STANDARD.CO.UK** | UK | 189,806 | 4,028 | 2.1 | 181,251 | 479 |
| **METRO.CO.UK** | UK | 170,989 | 105 | 0 | 165,381 | 86 |
| **WALESONLINE.CO.UK** | UK | 168,752 | 351 | 0.2 | 98,600 | 373 |
| **NEWS.SKY.COM** | UK | 86,114 | 1,568 | 1.8 | 80,694 | 79 |
| **SCOTSMAN.COM** | UK | 66,818 | 1,739 | 2.6 | 44,823 | 125 |
| **ITV.COM** | UK | 63,426 | 119 | 0.1 | 58,714 | 84 |
| **ECONOMIST.COM** | UK | 24,006 | 25 | 0.1 | 18,314 | 81 |
| **MORNINGSTARONLINE.CO.UK** | UK | 14,371 | 435 | 3 | 13,636 | 104 |
| **CHANNEL4.COM** | UK | 8,876 | 45 | 0.5 | 8,607 | 81 |
| **TOTAL** | UK | 7,081,296 | - | - | 4,977,792 | - |
| **BREITBART.COM** | USA | 473,460 | 68 | 0 | 175,292 | 696 |
| **LATIMES.COM** | USA | 428,327 | 477 | 0.1 | 228,673 | 3,911 |
| **WSJ.COM** | USA | 428,115 | 333 | 0 | 177,647 | 254 |
| **FOXNEWS.COM** | USA | 418,674 | 3,988 | 0.9 | 289,504 | 321 |
| **WASHINGTONPOST.COM** | USA | 375,562 | 2,901 | 0.7 | 292,075 | 431 |
| **UPI.COM** | USA | 370,826 | 27 | 0 | 138,082 | 72 |
| **CHICAGOTRIBUNE.COM** | USA | 291,095 | 252 | 0 | 146,505 | 1,948 |
| **NEWS.YAHOO.COM** | USA | 242,964 | 9,886 | 4 | 223,051 | 224 |

| | | | | | | |
|---|---|---|---|---|---|---|
| **USATODAY.COM** | USA | 238,906 | 1,619 | 0.6 | 231,819 | 113 |
| **CBSNEWS.COM** | USA | 216,257 | 4,809 | 2.2 | 193,922 | 1,876 |
| **BOSTONGLOBE.COM** | USA | 206,029 | 3,119 | 1.5 | 177,633 | 135 |
| **NYPOST.COM** | USA | 201,149 | 36 | 0 | 198,003 | 62 |
| **ABCNEWS.GO.COM** | USA | 200,014 | 3,050 | 1.5 | 172,410 | 223 |
| **NYTIMES.COM** | USA | 197,711 | 387 | 0.1 | 155,450 | 187 |
| **CNBC.COM** | USA | 191,100 | 1,795 | 0.9 | 168,541 | 509 |
| **THEHILL.COM** | USA | 171,544 | 138 | 0 | 170,377 | 222 |
| **BUSINESSINSIDER.COM** | USA | 145,345 | 956 | 0.6 | 117,468 | 1,056 |
| **NBCNEWS.COM** | USA | 140,918 | 266 | 0.1 | 126,075 | 219 |
| **WASHINGTONTIMES.COM** | USA | 139,276 | 112 | 0 | 114,203 | 292 |
| **WND.COM** | USA | 114,852 | 381 | 0.3 | 78,710 | 589 |
| **TIME.COM** | USA | 109,595 | 3,764 | 3.4 | 94,575 | 213 |
| **NEWSWEEK.COM** | USA | 103,173 | 236 | 0.2 | 101,583 | 493 |
| **EDITION.CNN.COM** | USA | 94,283 | 2 | 0 | 93,871 | 73 |
| **APNEWS.COM** | USA | 94,259 | 73 | 0 | 92,507 | 115 |
| **SLATE.COM** | USA | 87,595 | 562 | 0.6 | 64,312 | 213 |
| **POLITICO.COM** | USA | 83,675 | 320 | 0.3 | 79,311 | 639 |

| | | | | | | |
|---|---|---|---|---|---|---|
| **NPR.ORG** | USA | 70,753 | 827 | 1.1 | 61,537 | 265 |
| **MSNBC.COM** | USA | 54,395 | 17 | 0 | 53,084 | 55 |
| **THEATLANTIC.COM** | USA | 49,676 | 25 | 0 | 46,422 | 182 |
| **HUFFPOST.COM** | USA | 38,524 | 1,602 | 4.1 | 36,119 | 286 |
| **CSMONITOR.COM** | USA | 37,194 | 33 | 0 | 28,495 | 148 |
| **WESTERNJOURNAL.COM** | USA | 34,981 | 686 | 1.9 | 31,682 | 106 |
| **INQUIRER.COM** | USA | 31,029 | 43 | 0.1 | 29,445 | 229 |
| **TOTAL** | USA | 6,081,256 | - | - | 4,388,383 | - |
| **FRANCETVINFO.FR** | France | 694,187 | 726 | 0.1 | 573,917 | 2,675 |
| **LEFIGARO.FR** | France | 507,333 | 469 | 0 | 291,187 | 718 |
| **LEMONDE.FR** | France | 288,325 | 3,587 | 1.2 | 203,071 | 1,208 |
| **LEXPRESS.FR** | France | 239,065 | 143 | 0 | 235,163 | 300 |
| **20MINUTES.FR** | France | 207,226 | 9,230 | 4.4 | 167,383 | 574 |
| **LEPOINT.FR** | France | 177,423 | 19 | 0 | 171,828 | 183 |
| **LIBERATION.FR** | France | 166,487 | 2,002 | 1.2 | 149,082 | 207 |
| **LCI.FR** | France | 113,084 | 221 | 0.1 | 94,273 | 155 |
| **BFMTV.COM** | France | 66,531 | 119 | 0.1 | 65,704 | 216 |
| **TOTAL** | France | 2,459,661 | - | - | 1,951,608 | - |

| | | | | | | |
|---|---|---|---|---|---|---|
| **WELT.DE** | Germany | 362,621 | 980 | 0.2 | 307,070 | 342 |
| **SPIEGEL.DE** | Germany | 313,438 | 2,247 | 0.7 | 270,326 | 1,749 |
| **FOCUS.DE** | Germany | 276,482 | 8,907 | 3.2 | 250,533 | 351 |
| **FAZ.NET** | Germany | 266,962 | 929 | 0.3 | 215,398 | 2,891 |
| **N-TV.DE** | Germany | 251,589 | 1,506 | 0.5 | 239,727 | 979 |
| **HANDELSBLATT.COM** | Germany | 221,696 | 5,628 | 2.5 | 139,412 | 391 |
| **BILD.DE** | Germany | 194,428 | 570 | 0.2 | 187,926 | 610 |
| **STERN.DE** | Germany | 174,504 | 5,381 | 3 | 115,613 | 4,529 |
| **TAGESSPIEGEL.DE** | Germany | 155,218 | 130 | 0 | 133,925 | 293 |
| **NOZ.DE** | Germany | 138,794 | 509 | 0.3 | 84,747 | 447 |
| **FR.DE** | Germany | 117,738 | 4,856 | 4.1 | 64,094 | 145 |
| **ABENDBLATT.DE** | Germany | 109,148 | 5,794 | 5.3 | 98,415 | 170 |
| **TAGESSCHAU.DE** | Germany | 104,377 | 3,021 | 2.8 | 50,985 | 88 |
| **MORGENPOST.DE** | Germany | 100,603 | 6,317 | 6.2 | 87,741 | 188 |
| **BZ-BERLIN.DE** | Germany | 55,698 | 202 | 0.3 | 54,690 | 250 |
| **NDR.DE** | Germany | 50,989 | 10 | 0 | 10,640 | 93 |
| **WDR.DE** | Germany | 40,966 | 1,306 | 3.1 | 18,667 | 332 |
| **RTL.DE** | Germany | 20,105 | 29 | 0.1 | 18,494 | 77 |

| | | | | | | |
|---|---|---|---|---|---|---|
| **TOTAL** | Germany | 2,955,356 | - | - | 2,348,403 | - |
| **THESUN.IE** | Ireland | 196,122 | 522 | 0.2 | 161,165 | 29 |
| **INDEPENDENT.IE** | Ireland | 191,800 | 5,250 | 2.7 | 179,924 | 577 |
| **THEJOURNAL.IE** | Ireland | 178,462 | 222 | 0.1 | 102,461 | 72 |
| **IRISHTIMES.COM** | Ireland | 176,304 | 103 | 0 | 161,235 | 393 |
| **IRISHEXAMINER.COM** | Ireland | 140,383 | 7,523 | 5.3 | 126,892 | 282 |
| **IRISHMIRROR.IE** | Ireland | 104,582 | 311 | 0.2 | 66,671 | 64 |
| **BREAKINGNEWS.IE** | Ireland | 78,338 | 234 | 0.2 | 76,423 | 82 |
| **RTE.IE** | Ireland | 32,530 | 51 | 0.1 | 30,827 | 127 |
| **TOTAL** | Ireland | 1,098,521 | - | - | 905,598 | - |
| **EURONEWS.COM** | International | 138,093 | 4,553 | 3.2 | 129,879 | 168 |
| **DW.COM** | International | 112,242 | 2,599 | 2.3 | 100,029 | 278 |
| **RT.COM** | International | 103,067 | 1,574 | 1.5 | 99,282 | 63 |
| **ALJAZEERA.COM** | International | 66,785 | 34 | 0 | 65,477 | 458 |
| **FRANCE24.COM** | International | 58,714 | 710 | 1.2 | 57,005 | 49 |
| **RFI.FR** | International | 11,550 | 42 | 0.3 | 11,317 | 34 |
| **TOTAL** | International | 490,451 | - | - | 462,989 | - |

| | | | | | | |
|---|---|---|---|---|---|---|
| **STUFF.CO.NZ** | New Zealand | 276,231 | 22,793 | 8.2 | 221,823 | 641 |
| **NZHERALD.CO.NZ** | New Zealand | 274,618 | 3,703 | 1.3 | 245,787 | 515 |
| **SCOOP.CO.NZ** | New Zealand | 124,309 | 88 | 0 | 119,624 | 708 |
| **NEWSHUB.CO.NZ** | New Zealand | 49,696 | 2,141 | 4.3 | 46,817 | 95 |
| **RNZ.CO.NZ** | New Zealand | 18,944 | 1,041 | 5.4 | 16,999 | 77 |
| **TOTAL** | New Zealand | 743,798 | - | - | 651,050 | - |
| **RBC.RU** | Russia | 382,762 | 751 | 0.1 | 246,376 | 255 |
| **RIA.RU** | Russia | 352,608 | 62 | 0 | 282,422 | 206 |
| **KOMMERSANT.RU** | Russia | 318,046 | 314 | 0 | 232,298 | 289 |
| **GAZETA.RU** | Russia | 283,339 | 80 | 0 | 240,404 | 566 |
| **LENTA.RU** | Russia | 267,014 | 508 | 0.1 | 263,490 | 284 |
| **INTERFAX.RU** | Russia | 252,237 | 194 | 0 | 228,483 | 404 |
| **RAMBLER.RU** | Russia | 245,294 | 1,622 | 0.6 | 206,648 | 338 |
| **MK.RU** | Russia | 245,201 | 230 | 0 | 239,912 | 453 |
| **NTV.RU** | Russia | 224,220 | 377 | 0.1 | 136,400 | 797 |
| **AIF.RU** | Russia | 216,288 | 21 | 0 | 172,920 | 345 |

| | | | | | | |
|---|---|---|---|---|---|---|
| **VESTI.RU** | Russia | 188,944 | 1,527 | 0.8 | 168,850 | 200 |
| **IZ.RU** | Russia | 179,549 | 15 | 0 | 165,034 | 346 |
| **KP.RU** | Russia | 157,505 | 297 | 0.1 | 152,472 | 381 |
| **TASS.RU** | Russia | 153,497 | 945 | 0.6 | 151,541 | 574 |
| **RG.RU** | Russia | 142,451 | 75 | 0 | 139,365 | 279 |
| **NEWSRU.COM** | Russia | 105,902 | 496 | 0.4 | 100,704 | 53 |
| **TVZVEZDA.RU** | Russia | 90,639 | 4,364 | 4.8 | 84,260 | 348 |
| **NG.RU** | Russia | 86,974 | 3,341 | 3.8 | 66,217 | 329 |
| **REN.TV** | Russia | 72,092 | 100 | 0.1 | 71,029 | 662 |
| **TOTAL** | Russia | 3,964,562 | - | - | 3,348,825 | - |
| **ABC.ES** | Spain | 653,473 | 2,448 | 0.3 | 330,934 | 3,378 |
| **COPE.ES** | Spain | 419,512 | 3,678 | 0.8 | 242,826 | 3,332 |
| **20MINUTOS.ES** | Spain | 404,122 | 1,535 | 0.3 | 248,585 | 762 |
| **ELMUNDO.ES** | Spain | 355,466 | 636 | 0.1 | 201,989 | 1,198 |
| **ELPAIS.COM** | Spain | 311,945 | 350 | 0.1 | 200,784 | 636 |
| **CADENASER.COM** | Spain | 297,416 | 1,224 | 0.4 | 251,223 | 9,043 |
| **ELPERIODICO.COM** | Spain | 292,584 | 1,251 | 0.4 | 221,135 | 240 |
| **EUROPAPRESS.ES** | Spain | 287,886 | 5,235 | 1.8 | 276,158 | 1,296 |

| | | | | | | |
|---|---|---|---|---|---|---|
| **LARAZON.ES** | Spain | 268,640 | 7,565 | 2.8 | 166,949 | 304 |
| **ELESPANOL.COM** | Spain | 231,377 | 4,429 | 1.9 | 137,183 | 1,668 |
| **LAVANGUARDIA.COM** | Spain | 199,272 | 681 | 0.3 | 194,336 | 1,508 |
| **HUFFINGTONPOST.ES** | Spain | 190,885 | 629 | 0.3 | 109,330 | 1,844 |
| **ELDIARIO.ES** | Spain | 165,130 | 405 | 0.2 | 124,094 | 3,222 |
| **RTVE.ES** | Spain | 132,005 | 361 | 0.2 | 108,166 | 103 |
| **PUBLICO.ES** | Spain | 131,274 | 335 | 0.2 | 111,370 | 4,921 |
| **TELECINCO.ES** | Spain | 64,818 | 2,849 | 4.3 | 58,657 | 201 |
| **ANTENA3.COM** | Spain | 52,484 | 132 | 0.2 | 49,916 | 236 |
| **ONDACERO.ES** | Spain | 50,935 | 88 | 0.1 | 49,604 | 196 |
| **CUATRO.COM** | Spain | 42,919 | 738 | 1.7 | 39,472 | 413 |
| **TOTAL** | Spain | 4,552,143 | - | - | 3,122,711 | - |
| **TOTAL** | Overall | 34,277,756 | - | - | 26,077,939 | - |

### Supplementary Section 2. Coverage.

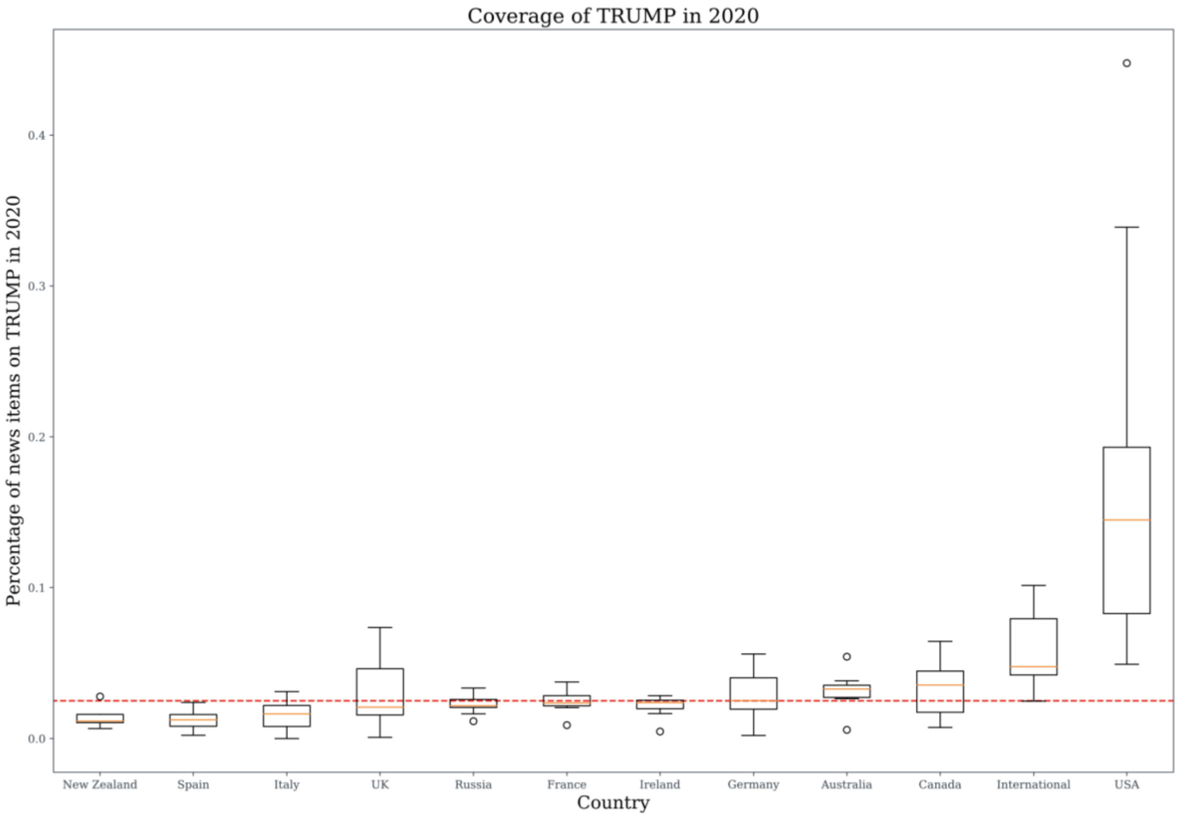


**Supplementary Figure 1.** Volume of coverage of Donald Trump in 2020.

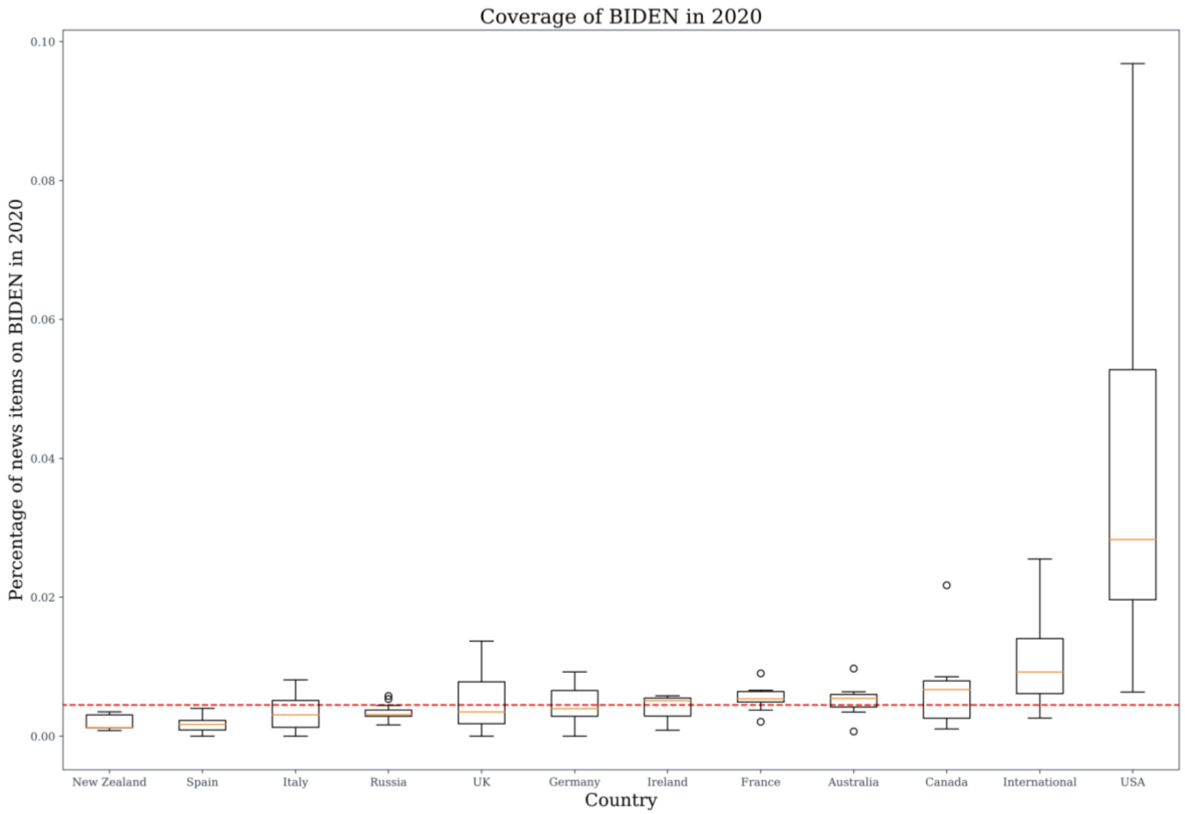


**Supplementary Figure 2.** Volume of coverage of Joe Biden in 2020.

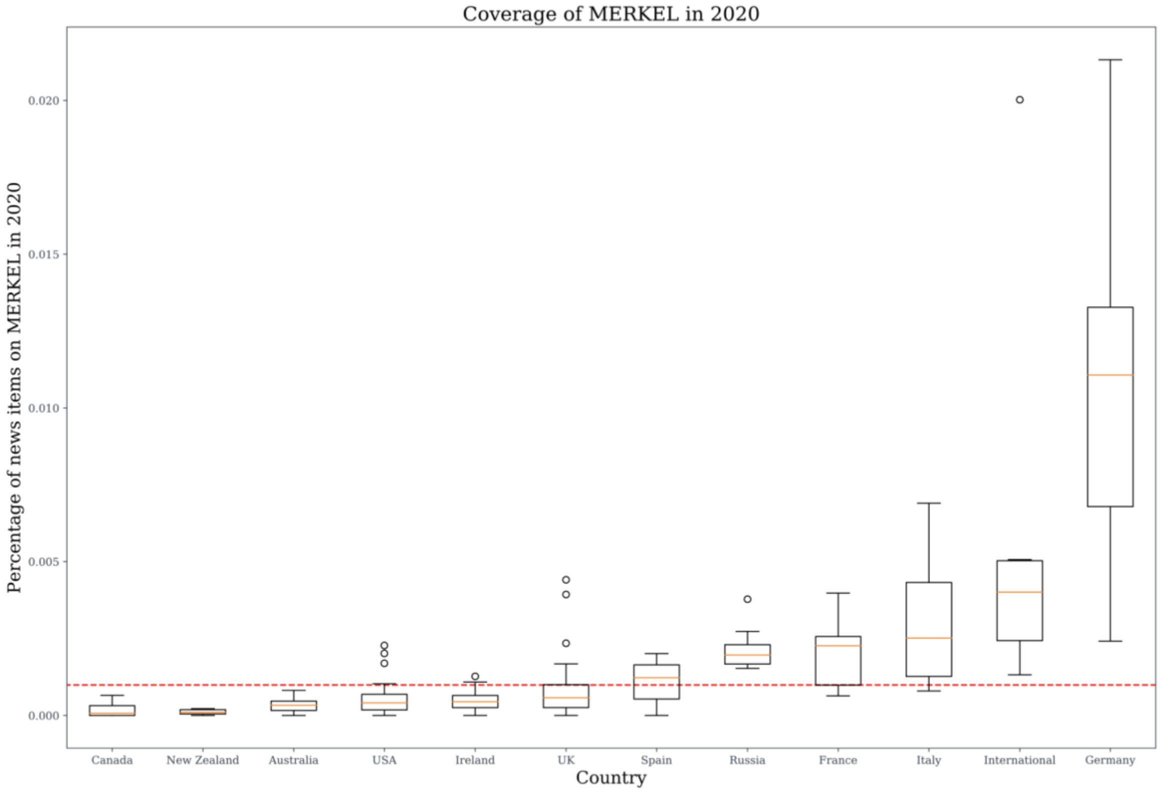


**Supplementary Figure 3.** Volume of coverage of Angela Merkel in 2020.

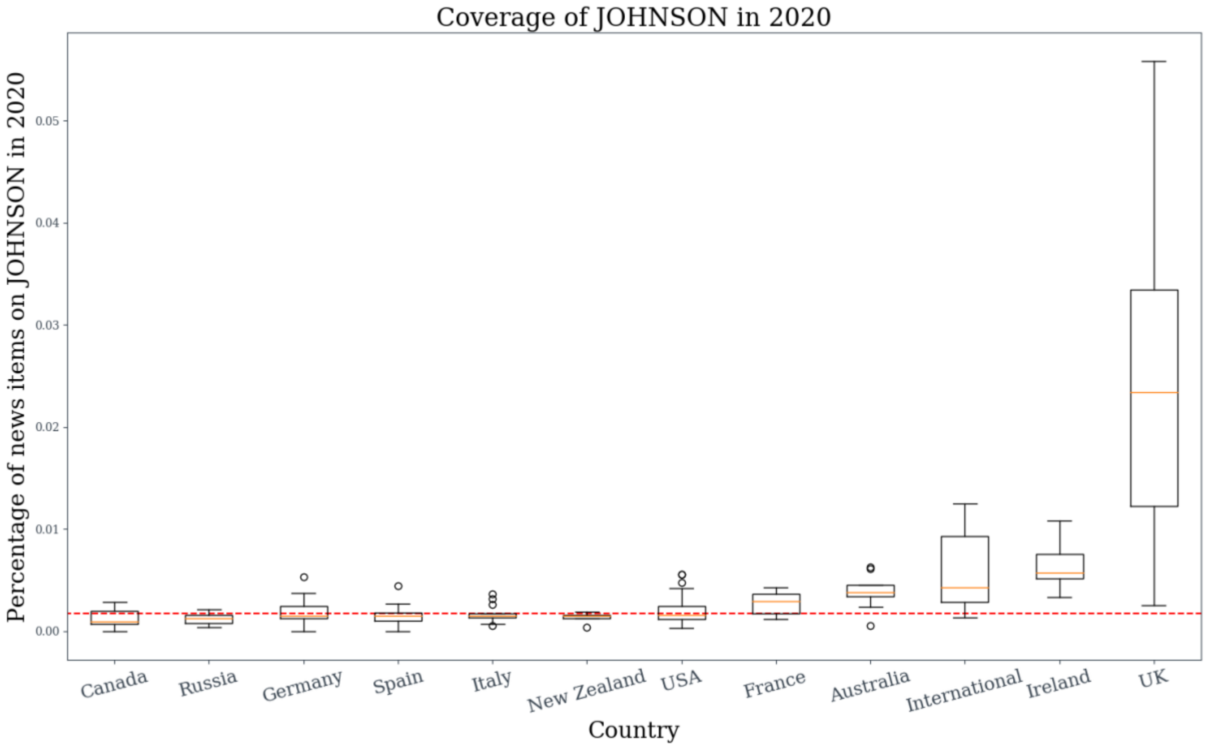


**Supplementary Figure 4.** Volume of coverage of Boris Johnson in 2020.

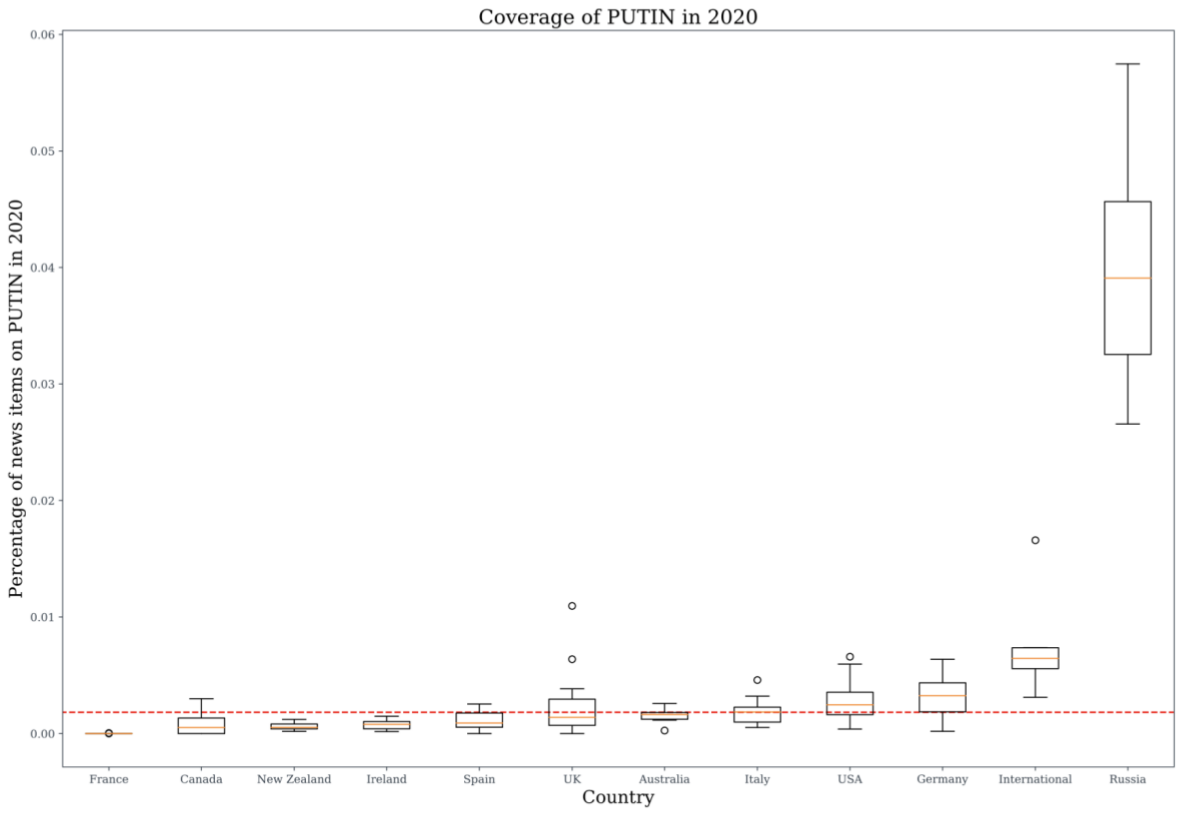


**Supplementary Figure 5.** Volume of coverage of Vladimir Putin in 2020.

### Supplementary Section 3. Sentiment.

Topics such as *CANCER* however, an order of magnitude more negative, receive much less coverage than COVID-19. Out of 2,149,585 front page articles from English-speaking ONSs in 2020 we identified 589,701 as *COVID* and 9,548 as *CANCER* (Table 3). The volume of the coverage for a given topic might play a role in being able to polarize the overall sentiment perception. To illustrate this, in supplementary Figure 6 we plotted the sentiment skew as in Figure 2, but with individual ONS sentiment differences weighted by the proportion of topic coverage. The sentiment skew becomes more pronounced for topics which received a large amount of media coverage (e.g. *COVID*) than for topics with less media coverage (e.g. *CANCER*). This intuitively illustrates the higher contribution to sentiment polarization for topics with relatively lower negativity but high coverage volume (e.g. *COVID*) than more negative topics that receive less coverage (e.g. *CANCER*).

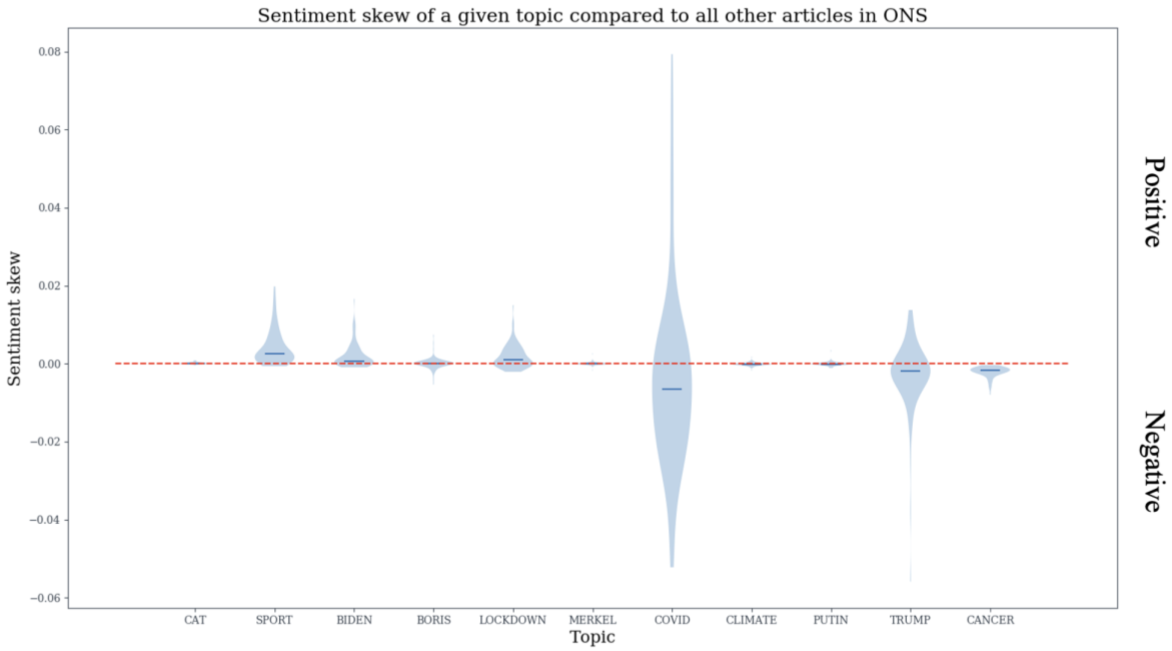


**Supplementary Figure 6.** Relative Sentiment Skew of news in 2020, weighted by percentage volume coverage in 2020. The rsskew formula was adjusted by the proportion of coverage of a given topic as $(\mu_{ONS,TOPIC} - \mu_{ONS,\underline{TOPIC}})$*$coverage_{ONS,TOPIC}$ where $coverage_{ONS,TOPIC}$ is the ratio of articles on a specific topic in 2020 in a specific ONS.

### Supplementary Section 4. COVID-19 Sub-topics.

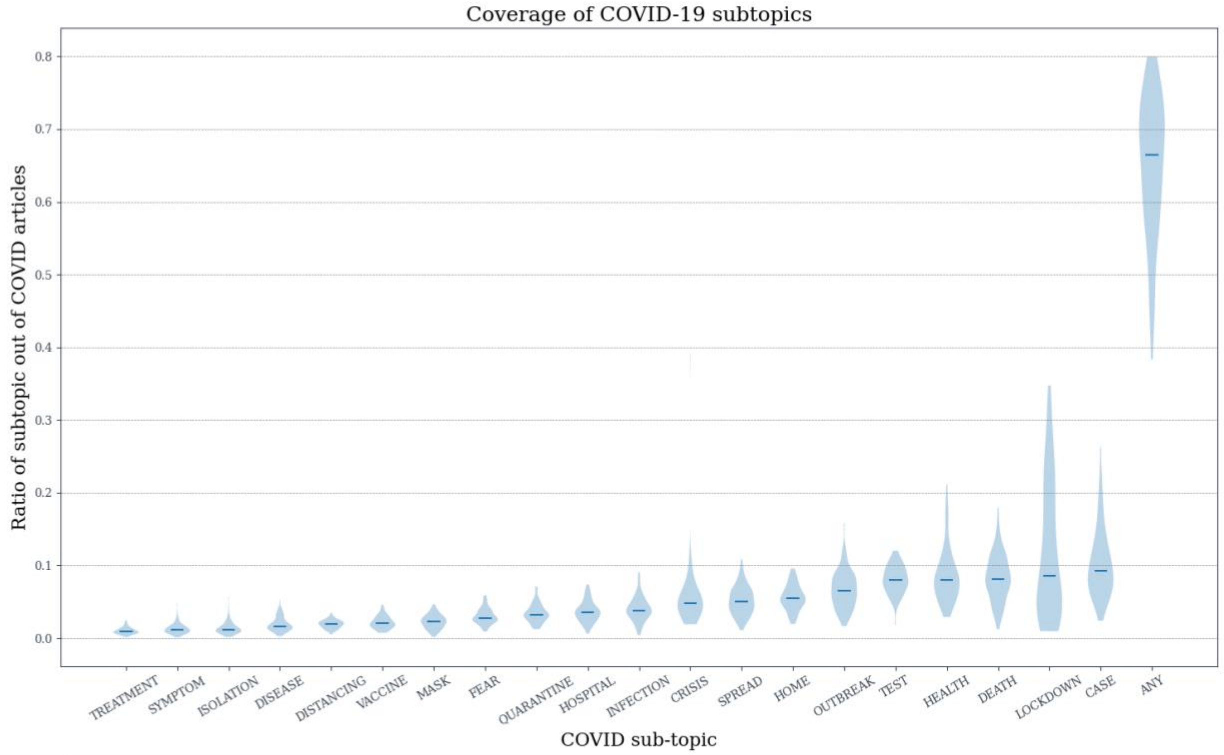


**Supplementary Figure 7. Subtopic coverage per ONS.** For each of 91 English-speaking ONS we calculated the proportion of all the COVID articles that a given subtopic accounted for. The violin plots are aggregates of such proportions per-subtopic from each of 91 ONS. The subtopic 'ANY' denotes the proportion of articles that can be classified as one of the other subtopics in the Figure.

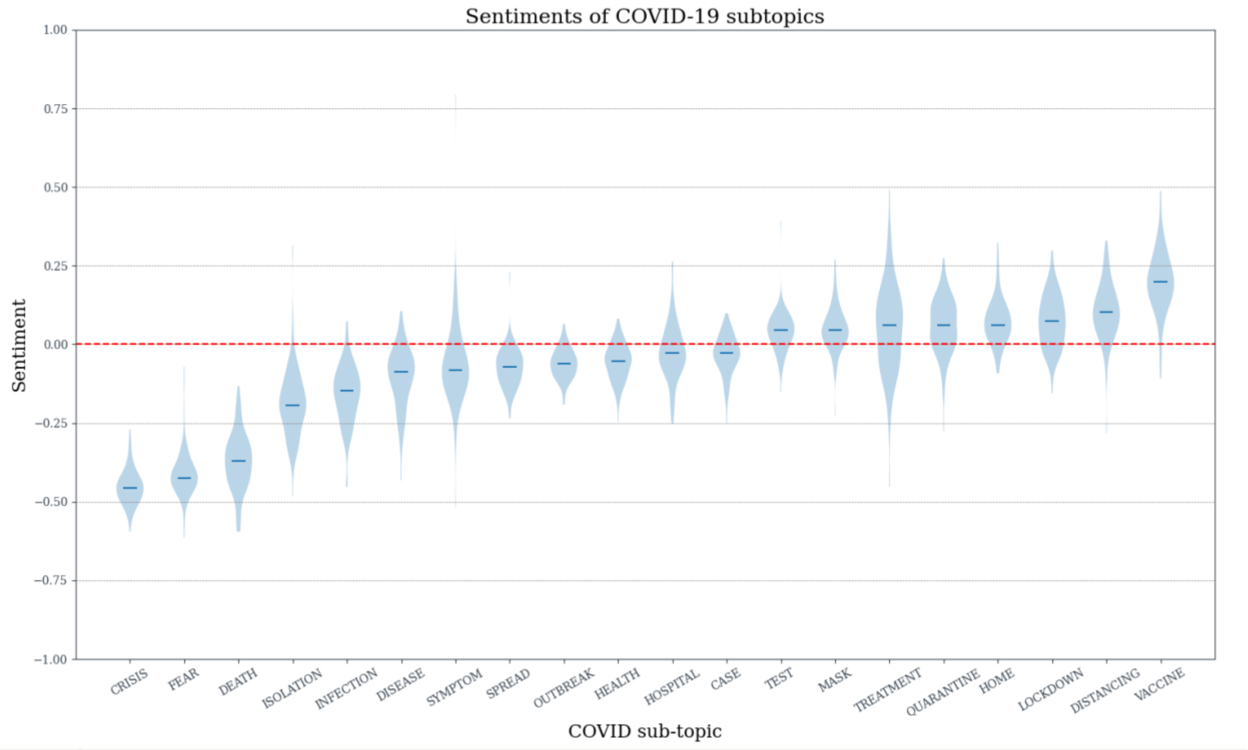


**Supplementary Figure 8. Subtopic sentiment per ONS.** For each of 91 English-speaking ONS we calculated the mean VADER sentiment of all the articles that a given subtopic accounted for. The violin plots are aggregates of such mean sentiments per-subtopic from each of 91 ONS.

### Supplementary Section 5. Estimating sentiment polarization after removing a number of articles.

We tested whether removing all articles identified as specific topics (e.g. COVID-19) from ONS would result in a statistically significant polarization towards either positive or negative sentiment. We noted the mean sentiment after removing all articles on a given topic from a specific ONS. We then assessed how extreme the given sentiment mean was by approximating its distribution by repeatedly removing the same number of random articles from the outlet.

Let ONS$_{all}$ denote all articles in a given ONS, $ONS_{\underline{TOPIC}}$ all articles not identified as topic in a given ONS and ONS$_{TOPIC}$ all articles on a given topic in the given ONS. We noted the mean sentiment of 2020 articles when a given topic was removed $\mu_{ONS,\underline{TOPIC}}$ . We then sampled |ONS$_{TOPIC}$| articles from ONS$_{all}$ and noted the sentiment of the remaining articles denoted $\mu_{ONS,sample}$ . Each such sample provided an indicator of possible average sentiment when the same number of articles as |ONS$_{TOPIC}$| is sampled. For each ONS the procedure of producing $\mu_{ONS,sample}$ was repeated min(1000,|ONS$_{TOPIC}$|) times, approximating the distribution of sentiment means upon removal of the given number of articles. We noted the number of times

$\mu_{ONS,\underline{TOPIC}}$ was higher than $\mu_{ONS,sample}$ as $H_{ONS}$ and the number where it was lower as $L_{ONS}$. Large $H_{ONS}$ value indicates that removing TOPIC articles from ONS on average results in sentiment polarization towards positive, as compared to random sampling the same number of articles from ONS and vice versa for $L_{ONS}$. In both cases we set the significance level at 5% ($H_{ONS}>0.95\%$ and $L_{ONS}<0.05$). Since we had 91 English-speaking ONSs, we corrected for multiple testing dividing by 91 (the Bonferroni correction). For each ONS and a given topic we therefore reported whether removing a topic had a significant polarization towards either positive or negative sentiment ($H_{ONS}>0.95/91$ and $L_{ONS}<0.05/91$) or whether no determination could be made ($H_{ONS}<0.95/91$ and $L_{ONS}>0.05/91$).
